# Requirements of a knowledge support tool for self-harm in general practice: a mixed-methods study with patients, carers, and clinicians

**DOI:** 10.64898/2026.11.14.26368111

**Authors:** Harriet Cant, Faraz Mughal, Sadika Asmal, Katharine Petersen, Jenny Groves, Emily Gardiner, Peter Bower, Roger T. Webb, Nav Kapur, Jodie Westhead, Victoria Palin, Alexander Pate, Tjeerd van Staa, Leah Quinlivan

## Abstract

**Background:** General practice has a central role in care following self-harm. However, clinicians frequently report uncertainty about best practice and patients describe poor care. A knowledge support tool is an add-on to electronic health systems that may improve services by providing personalised care information.

**Aim:** To understand what general practice clinicians, patients and carers need from a novel knowledge support tool for self-harm.

**Design and Setting:** Mixed-methods, using quantitative ratings, focus groups and follow-on interviews with general practice clinicians and patients/carers with lived experience of self-harm.

**Method:** Clinicians were recruited through social media and professional networks, and patients/carers through a third-sector organisation. During four focus groups and follow-on interviews, we explored the utility of potential content and functionality of a knowledge support tool. Quantitative utility ratings were summarised descriptively and qualitative data were analysed thematically using the framework method. People with lived/living experience were involved throughout.

**Results:** 13 clinicians and 14 patients/carers participated. Requirements of the knowledge support tool were captured across three qualitative themes: facilitating person-centred care, accessibility and usability, and balancing competing priorities of general practice. Across these themes, patients’ and carers’ needs were centred on promoting compassionate relationships and patient agency. Clinicians shared these needs but balanced them with a need to manage patient expectations and practice defensively under limited resources.

**Conclusion:** Our findings inform the core design of a knowledge support tool and suggest that such a tool could support the assessment and management of self-harm in general practice.

**How this fits in:** General practice plays a key role in providing care for people who have self-harmed, yet clinicians report feeling underconfident in managing patients, who themselves frequently describe poor experiences of care. Research suggests that reducing knowledge gaps amongst clinicians, patients and carers may improve patient safety and experiences of help-seeking in this setting, where practical guidance for clinicians is currently limited. A ‘knowledge support tool’ is a novel intervention that can be used during consultations to provide timely and personalised information that supports high-quality caregiving; however, less is known about how such a tool should be designed for general practice self-harm care. We outline what general practice clinicians and people with lived experience of self-harm need from a knowledge support tool for self-harm.

## Introduction

Self-harm, defined as intentional self-poisoning or injury irrespective of suicidal intent, is a major public health concern and lifetime prevalence rates have more than doubled over the past two decades.(1) Although there are multiple reasons for engaging in self-harm, people who have self-harmed are many times more likely to subsequently die by suicide than those who have not.(2) National guidance recommends that a specialist-led psychosocial assessment is promptly provided following a self-harm episode.(3) However, access to mental health care is limited and referral rates are low, resulting in general practice being a key healthcare setting for people who have self-harmed.(4, 5)

Despite the central role played by general practice, many general practitioners (GPs) are unaware of clinical guidelines for self-harm, which may result in unsafe practice.(4, 6) Both GPs and practice nurses have reported a lack of confidence in assessing and talking to people who have self-harmed.(7, 8) Suicide prevention strategies emphasise the importance of assessing individual needs, risks, and the use of collaboratively developed management plans,(3, 9, 10) but clinicians have reported difficulties in conducting thorough assessments during time pressured appointments.(11) The ability to review a patient’s medical history to inform care is hindered by inconsistent coding practices and the poor usability and interoperability of healthcare record systems.(8, 12, 13) Patients are often unaware of which services they can access outside of general practice, or may not be offered information about any.(14, 15)

Interventions to reduce knowledge gaps amongst clinicians, patients and carers may improve patient safety and the quality of general practice services for self-harm, including provision of tailored assessment support and information about medicines and signposting.(9, 16–18) Currently, there is limited research into interventions to support the management of self-harm in general practice.

A digital knowledge support tool aims to provide clinicians, patients and carers with personalised information to optimise caregiving, and are distinguished from decision support tools through the absence of clinical recommendations. The tool is displayed on a clinician’s computer and may be used during or outside of consultations with a patient. The knowledge support tool acts as an add-on to general practice electronic health systems, enabling it to extract relevant information directly from the patients’ records to personalise content. Information entered into or generated by the tool may be automatically coded back to patients’ records. A recent study (Building Rapid Interventions to improve Therapies; BRIT2) developed a knowledge support tool for common infections that includes contextualised risk information, evidence-based guidance and personalised patient leaflets. The BRIT2 tool is being evaluated as part of a cluster randomised controlled trial. Electronic health record templates are commonly used in general practice to support disease management but typically provide standardised rather than personalised information, limiting their ability to support care that is tailored to individual risks and needs. There is no template specifically designed to support the management of self-harm.

Given the novelty of a knowledge support tool for self-harm, understanding users’ needs is required to inform its development. Stakeholder involvement throughout the development process, described as ‘co-design’ in this study, can help to ensure that interventions are person-centred, contextually appropriate, equitable and utilised.(19, 20) Thus, we aimed to understand what general practice clinicians and people with lived experience of self-harm (patients and carers) need from the functionality and content of a knowledge support tool for self-harm.

## Methods

### Study design

We applied a mixed-methods design within a pragmatic paradigm,(21) employing focus groups and follow-on interviews with general practice clinicians and people with lived experience of self-harm. Focus groups introduced the concept of a knowledge support tool and involved participants collectively exploring suggested content and functionality of the tool through structured utility questions and group discussion.(22) Follow-on interviews aimed to validate and expand on focus group discussions.

### Patient and community involvement and engagement

People with experiences of attending healthcare services with self-harm and/or suicidal behaviour and/or of caring for people with such experiences were involved throughout all aspects of the study, including development of the grant application, design, topic guides, recruitment, analyses and dissemination. Healthcare professional contributors also informed the study design and guided interpretation of its findings. Three public contributors and one healthcare professional contributor are co-authors of this paper. A plain English summary of this study is provided in Supplementary Information S1.

### Participants and recruitment

Participants included general practice clinicians and people with lived experience of self-harm. Clinicians were defined as National Health Service (NHS) professionals from any clinical role who currently or previously have been involved in the assessment and/or management of patients who have self-harmed in general practice. Staff in non-clinical, specialist mental health or trainee roles were ineligible. Lived experience participants were aged 18 years or older with current or lifetime history of self-harm and experience of attending general practice services, or experience of caring for someone with these experiences. Lived experience participants will hereafter be referred to as ‘patients/carers’. Participants were also required to understand written and verbal English and have capacity to provide informed consent. Two focus groups were recruited for patients/carers, and two focus groups were recruited for clinicians. Focus groups with patients/carers and clinicians were held separately to encourage open discussion. Seven to eight participants were recruited for each focus group. Our sample size was based on ensuring data adequacy and information richness and was guided by previous research findings on this topic.(23–25)

Staff were recruited through social media and professional networks covering the North West and Midlands NHS regions in England. Some participants shared study information with colleagues (snowball sampling). Patients/carers were recruited purposively through collaboration with a UK-based third-sector organisation for people affected by self-harm. Individuals were invited to register their interest in participating by email. As a token of appreciation, participants were given shopping vouchers (£25 per hour of data collection) and participation certificates. Further methodological details are provided in Supplementary Information S2.

### Data collection procedures and tools

During May to August 202, clinicians participated online to accommodate ongoing clinical work, and patients/carers participated online or in-person at a community location, according to their preference. Semi-structured interviews (topic guides in Supplementary Information S3) were audio (in-person) or video (online, using Microsoft Teams) recorded using encrypted devices. All focus groups and interviews were facilitated by a researcher with experience in digital tool development (HC; White British, female) and a mental health services researcher (LQ; White Irish, female). Our approach to data collection was centred around inclusivity, accessibility and wellbeing (Supplementary Information S4).

#### Stage 1: Focus groups

Each focus group lasted two hours. Participants anonymously completed a demographic survey (electronically via Qualtrics or using pen-and-paper). All demographic data were self-reported. Participants explored the utility of suggested content and functionality of a knowledge support tool. Broad *types* of content and functionality were considered as well as *examples* which described how these may be delivered in practice. For example, one *type* of content was ‘information on support that may be available outside of appointments’, with ‘information on helplines’ provided as an *example*.

Participants individually rated the perceived utility of suggested content and functionality using a 10-point Likert scale, following an approach used in previous research to develop a knowledge support tool for common infections.(24) Ratings were collected anonymously using an online surveying tool (26) or pen-and-paper. For online focus groups, anonymous ratings were shared and semi-structured prompts explored the reasons for these ratings. Participants were encouraged to raise suggestions or requirements for a knowledge support tool outside of those presented within the topic guide.

#### Stage 2: Follow-on interviews

Interviews lasted for up to one hour and were conducted within nine weeks of the focus group (median of five weeks). Interviews began with a visual summary of focus group discussions to prompt reflection (Supplementary Figures S1-S2). Participants were asked semi-structured questions around the acceptability and implementation of the tool, during which they were prompted to further consider its content and functionality requirements. Findings around acceptability and implementation of the knowledge support tool will be reported separately.

#### Development of suggested content and functionality

Suggested content and functionality for the knowledge support tool were informed through input from the research team, healthcare professional and public contributors, findings from developing a knowledge support tool for common infections,(24, 27) and literature on best-practice care and opportunities to improve services for people who have self-harmed.(3, 4, 6–8, 11, 18, 28–30) The content and functionality examples were tailored for clinicians and patients/carers to ensure relevance and enhance understanding.

### Data analysis

Quantitative utility ratings were summarised descriptively using medians and their interquartile ranges.

Following transcription by a professional service, the framework method was used to analyse qualitative data (anonymised transcripts and hand-written notes).(31) The purpose of this approach was to understand “what works” in a knowledge support tool. The trustworthiness of the qualitative analysis was guided by Lincoln and Guba’s criteria (Supplementary Table S1).(32, 33)

Qualitative data analysis followed five stages as per Gale et al.(31) *Step 1:* researchers (HC and LQ) re-familiarised themselves with the data. *Step 2*: A transcript was coded in vivo (HC). Reflexive memos were made to document emergent patterns. Codes were reviewed and developed into an initial framework (HC, LQ), which grouped in vivo codes based on mixed inductive and deductive (using the topic guide) reasoning. *Step 3*: The framework was applied to subsequent transcripts (HC) and underwent iterative refinement to accommodate new insights (HC, LQ). The framework was independently reviewed against all transcripts (LQ) to assess consistency of application across researchers, which led to final framework refinements (see Supplementary Table S2). *Step 4:* Data were charted into matrices (HC) according to the framework. Using the matrices, domain summaries were formed. The matrices and summaries were independently reviewed against the transcripts (LQ). *Step 5:* The framework matrices and quantitative ratings were jointly interpreted with the multidisciplinary team (HC, LQ, FM, TvS, NK, RW, PB, SA, KP) and mapped to themes that pertained to the study’s aims. Resulting themes were iteratively discussed and revised through team meetings and sharing of draft results, including with the wider authorship team (JG, EG, AP, VP) and with healthcare professional and public contributors.

Quantitative ratings were triangulated with qualitative findings to contextualise and inform the themes (*Step* 5 above; Supplementary Table S3). Data were analysed from May 2025 to May 2026. Microsoft Excel and R (V 4.5.1) were used for management and analysis of the quantitative data and NVivo Version 15 for qualitative data.(34, 35) This study has been reported in accordance with qualitative and mixed-methods research standards (Supplementary Tables S4-S6).(36–38)

## Results

In total, 14 patients/carers and 13 general practice clinicians took part in a focus group (Supplementary Figure S3). Characteristics of focus group participants are reported in Table 1 and additional characteristics relating to clinician job roles are in Table 2. Of these, 12 patients/carers and 11 clinicians took part in a follow-on interview.

**Table 1:** Focus group participant characteristics (self-report)

| Characteristic | Patients/carers<br>(N=14) | General practice clinicians<br>(N=13) |
|---|---|---|
| Gender, N (%) |  |  |
| Female | 10 | 7 |
| Age (in years), median (IQR) | 36 (30-48) | 42 (40-50) |
| Ethnicity, N (%) |  |  |
| White | 12 | 10 |
| Asian or Asian British | 2 | 3 |
| Neurodivergent, N (%) |  |  |
| Yes | 9 | 1 |
| No | 4 | 12 |
| Prefer not to say | 1 |  |
| Employment status, N (%) |  |  |
| Employed full-time | 3 | NA |
| Employed part-time | 3<sup>1</sup> | NA |
| Employed voluntary | 1<sup>1</sup> | NA |
| Not accessing work due to sickness or disability | 5 | NA |
| Student | 2 | NA |
| Carer | 1 | NA |
| Most recent access of general practice services for self-harm |  |  |
| Within 6 months | 9 | NA |
| 6 months to ≤1 year ago | 3 | NA |
| 1 to ≤2 years ago | 1 | NA |
| Not responded | 1 | NA |

N=number of participants; IQR=interquartile range.
<sup>1</sup>One participant reported working both on a part-time and voluntary basis.
Statistics are reported to the nearest whole number.

**Table 2:** Additional demographics relating to job role of general practice clinicians who participated in a focus groups (self-report)

| Characteristic | N |
|---|---|
| Job role (self-described) |  |
| GP | 8 |
| GP partner | 2 |
| Senior clinical pharmacist | 1 |
| Mental health nurse and CBT therapist | 1 |
| Mental health pharmacist | 1 |
| Time spent in current role (in years) |  |
| <5 | 4 |
| 5-<10 | 3 |
| 10-<20 | 2 |
| 20+ | 3 |
| Area where most clinical time is spent working (by deprivation level) |  |
| In areas of low deprivation | 4 |
| In areas of mixed deprivation | 6 |
| In areas of high deprivation | 2 |
| Not responded | 1 |

N=number of participants, GP=general practitioner, CBT=cognitive behavioural therapy.

### Quantitative ratings of perceived utility for suggested content and functionality of the knowledge support tool

Participants provided utility ratings for suggested content and functionality of a knowledge support tool (Figure 1) and illustrative examples (Supplementary Table S7). Ratings are reported as median values (with their interquartile range).

**Figure 1:**
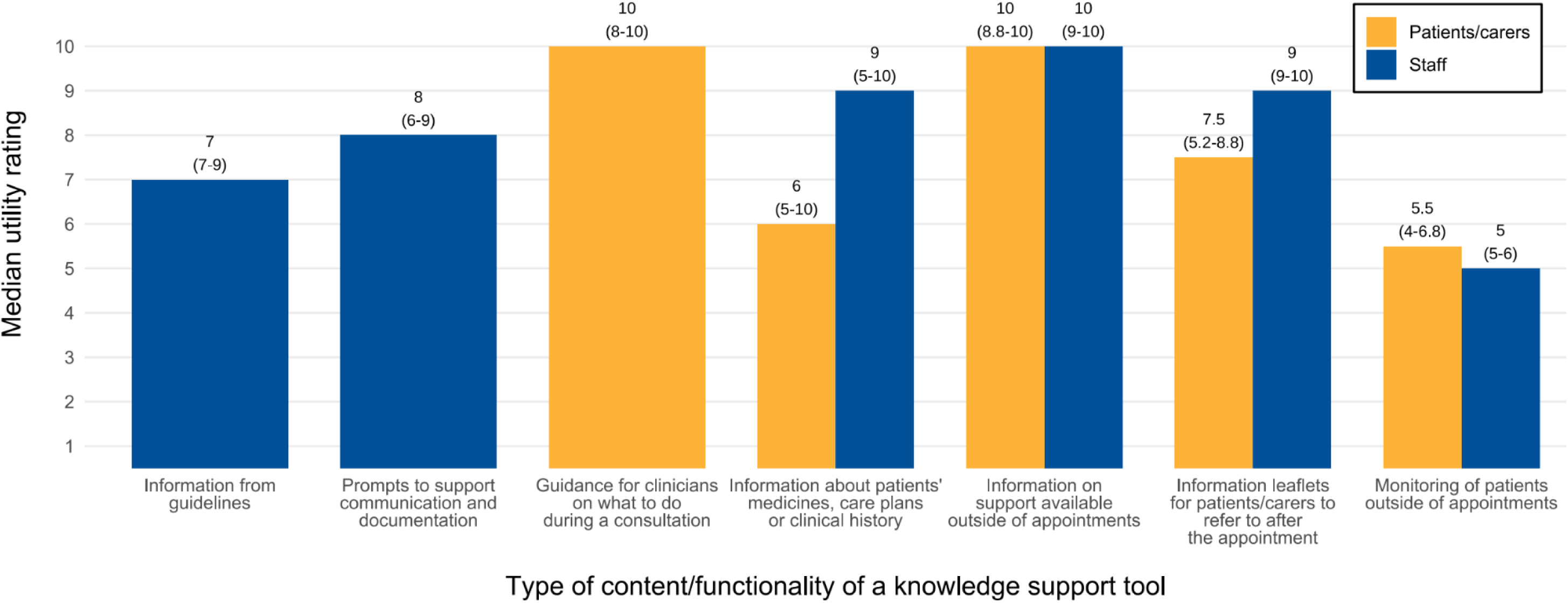
Median utility ratings for suggested content and functionality of a knowledge support too. Median values (with their interquartile ranges) are presented above each bar and have been reported to one decimal point. Utility was ranked on a scale of 1-10, with 1=completely not useful, 5=neutral, and 10=very useful. Some patients/carers submitted more than one response (one participant attended both focus groups). All responses were provided anonymously and therefore multiple responses could not be identified. The full results, including the number of responses for each item, are included in Supplementary Table S7. ‘Information from guidelines’ and ‘Prompts to support communication and documentation’ were presented only to clinicians, as these both pertain to healthcare professional facing content (e.g. it was not expected that patients or carers would use or be familiar with clinical guidelines). Patients/carers were instead presented with the broader option, ‘Guidance for clinicians on what to do during a consultation’, which was intended to broadly capture both best-practice guidance and support for communication. As such, direct comparisons between patients/carers and clinicians were not possible for these items. Item labels have been shortened for brevity; full wording is available in Supplementary Table S7.

Participants reported the highest utility for ‘information on support available outside of appointments’ (patients/carers: 10.0 [8.8-10]; clinicians: 10.0 [9.0-10.0]) and the lowest utility for ‘monitoring of patients outside of appointments’ (patients/carers: 5.5 [4.0-6.8]; clinicians: 5.0 [5.0-6.0]). The largest difference in ratings between the participant groups was for ‘information about a patient’s medicines, care plans or clinical history’, with patients/carers reporting this to be less useful than clinicians (patients/carers: 6.0 [5.0-10.0]; clinicians: 9.0 [5.0-10.0]).

Discussions following each rating exercise enabled participants to explore and explain the variation in responses.

### Qualitative themes: clinician, patient and carer needs for the functionality and content of a knowledge support tool

The needs of clinicians, patients and carers for a knowledge support tool were captured across three overarching themes (Figure 2): (1) ‘Facilitating person-centred care, (2) ‘Accessibility and usability’, and (3) ‘Balancing competing priorities of general practice. Illustrative verbatim quotes are presented in Supplementary Table S8, with selected quotes presented below.

**Figure 2:**
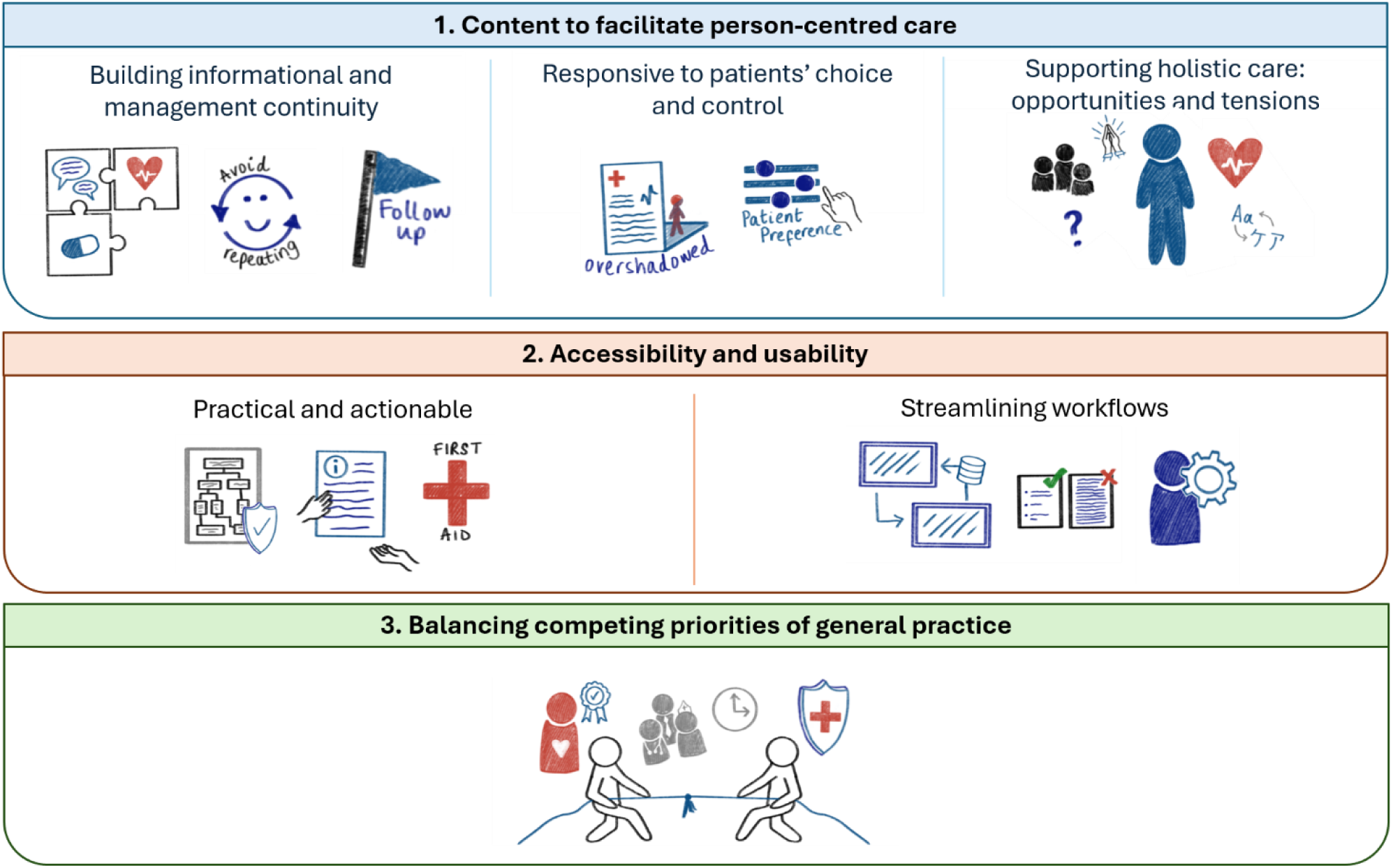
Visual representation of themes and subthemes of general practice clinicians, patient and carer requirements of a knowledge support tool for primary care self-harm services.

#### Theme 1: Content to facilitate person-centred care

##### Subtheme 1: Building informational and management continuity

Both clinicians and patients/carers valued summaries of hard-to-access information from patients’ health records. This was expected to improve understanding of patients’ histories whilst reducing burden on them to repeat painful experiences. Staff called for summaries to detail service use, care plans and contextual medication history. However, both clinician and patient/carer participants noted that such data are often buried within unstructured letters or not in general practice records altogether, particularly for patients with complex health and social needs.

> *“You have to trawl through the records to find out what’s gone on in the past, especially if they go from one doctor to another. So, having a summary of when they were prescribed as a snapshot would be really helpful.”* C_FG13 (clinician)

Some patients/carers additionally suggested that the tool may support management continuity by recording which support options have been previously been discussed, prompting clinicians to follow-up with ongoing care and avoid offering management options that patients are already aware of or have identified as being unhelpful.

> *“Say you’ve gone three times and one of those times [a support option] has already been suggested to you, could it come up as part of a tool like ‘it’s been suggested previously’ so you’re not going over old ground.”* PC_FG1 (patient/carer)

##### Subtheme 2: Responsive to patients’ choice and control

Whilst patients/carers valued summaries of patients’ records, they expressed concern that summarising historic diagnoses may cause clinicians to make assumptions about the cause of their self-harm, lead to exclusion from services, or enable potential misdiagnoses to influence care. Patients/carers therefore emphasised the need for patient agency over how the information within summaries is introduced and used during consultations. Similarly, patients/carers were concerned that functionality enabling patients to log their mood and thoughts outside consultations could reduce patients’ control over who can access or use this information. Compared with patients/carers, clinicians placed less emphasis on patient control over how information is used during consultations, although some clinicians suggested the use of pre-consultation surveys to incorporate patient-reported information into summaries.

> *“I have a physical disability, but quite often if they see mental health they’ll associate the two… That link is so common that it makes me nervous that if I went in for something specific and they had it all up on a screen that would be the first impression”* PC_FG4 (patient/carer)

> *“Sometimes your physical health will take absolutely precedence over your mental health, even though the two are completely inter-related.”* PC_FG14 (patient/carer)

##### Subtheme 3: Supporting holistic care: opportunities and tensions

Both clinicians and patients/carers described the factors influencing self-harm as complex and multi-faceted. Patients/carers welcomed prompts that support clinicians to holistically explore the meaning and reasons for self-harm with patients, seeing this as important for establishing patient expectations, building a compassionate clinician-patient relationship, and ensuring meaningful assessments.

> *“The tool could prompt the GPs to ask certain questions. ‘Would you like to change something about what you do?’, ‘how do you feel about your self-harm? Is it something that you’ve accepted, something you like, something you don’t like?’, ‘do you feel has got worse and this is why you’re here today?”* PC_FG1 (patient/carer)

Staff described assessments and support provision as involving complex clinical reasoning and a personalised approach to exploring social, cultural, and environmental factors. Despite this, several clinicians called for risk or severity scores to assess trends, automate care processes and optimise limited resources. Other clinicians perceived scores to promote impersonal or exclusionary care.

> *“It will be really useful in my role for risk stratification, especially for allocation of appointments… We have a mental health care coordinator and ultimately we’d love her to be able to decide the appointments without us… You’re asking someone who’s not clinical to make the decision, but if a tool could make the decision that might help”* C_I9 (clinician)

> *“[Risk scores] lose the human humanistic side of what we’re trying to achieve… As a GP, which is the nice thing about being a GP, you try to be holistic, you try to take the whole person. And [a score] could potentially just strip somebody of that personification.”* C_FG9 (clinician)

Both participant groups emphasised the importance of adapting the knowledge support tool content to meet patients’ cultural and language needs; for instance through translated patient leaflets and prompts to consider religion. Considering co-existing conditions and broader contextual factors was deemed important for understanding care needs and providing patient-centred support.

#### Theme 2: Accessibility and usability

##### Subtheme 1: Practical and actionable

Staff and patients/carers valued content to promote care aligned with up-to-date guidance from the National Institute for Health and Care Excellence (NICE) without the need to reference external sources. However, participants described NICE guidance as offering abstract general principles over practical instruction, and as being reliant on the availability of secondary care services. In addition to NICE guidance, both clinicians and patients called for content and functionality that support clear and implementable action. Staff additionally valued the ability to demonstrate actions to provide clinicians with medicolegal protection. For example, participants from both groups welcomed information on available support services, and for this information to be detailed in leaflets that patients and carers may refer to when needed. Patients/carers additionally welcomed leaflets that provide accessible and practical information about referral pathways (such as waiting times) and wound care.

> *“Practical information which you can share with patients so that they’ve got safety netting… Having a documented plan for if things were to deteriorate is quite helpful from a medicolegal point of view as much as anything else.”* C_I1 (clinician)

Both clinicians and patients/carers also viewed communication prompts (e.g. suggested discussion points or guidance on phrasing or language) as useful to directly support conversations around care needs and risk factors. Several participants emphasised that communication prompts should prioritise understanding patients’ lived experiences and building clinician-patient rapport over ease-of-documentation, and should therefore limit the use of tick-box formats (e.g. yes/no questions). Patients/carers called for prompts and leaflets to be co-designed with patients to ensure that they use appropriate language and promote patient-centredness.

> *“The prompts are supposed to be to engage risk but also to start building a relationship and building trust. It’s not about ticking boxes, are we trying to cover our backs or are they actually really trying to get through to this person?”* PC_FG8 (patient/carer)

##### Subtheme 2: Streamlining workffows

Both participant groups valued content that supports workflow efficiency. Staff and patients/carers called for content to be concise and accessible, avoiding lengthy paragraphs of text and using visuals where appropriate for ease-of-communication. Staff also called for dynamic and patient-personalised content, responding to both information from the patient record and information inputted during the consultation to reduce screen clutter.

> *“If it’s screeds of text, that’s useless in the consultation. I’m sure you’ve come across Ardens. Often their tools are dynamic, so if you tick boxes, it then tells you what to do. That kind of thing is really helpful”* C_FG1 (clinician)

Staff valued functionality to automate time-consuming administrative processes, such as aiding documentation of consultations and collating up-to-date details for support services. Some clinicians described potential for the tool to simplify complex referral processes by automatically drafting referral letters, thereby reducing the post-consultation administrative workload. As a high proportion of referrals are rejected, provision of clear referral criteria and automatic drafting of referral letters may help to reduce unnecessary associated resource wastage.

Staff called for the tool to integrate with systems and resources already used within a practice, such as messaging systems and patient leaflets, to avoid it acting as an additional standalone system that may complicate workflows and contribute to digital burnout. As workflows across clinicians may differ, the tool must allow for non-prescriptive use, enabling users to have flexibility in which content is accessed and in what order.

#### Theme 3: Balancing competing priorities of general practice

Staff accounts reflected competing priorities between providing compassionate, high-quality care and maintaining service capacity within the resource constraints of a non-specialist setting. Staff were therefore cautious of the tool promoting general practice as the central provider of ongoing care, perceiving this as increasing workload demands and patient expectations beyond what can feasibly be met. As such, clinicians expressed concerns about functionality that enables patients to self-report symptoms or distress between consultations, as this may place additional expectation on general practice clinicians to respond to them in real-time or carry medicolegal implications if they do not. Some clinicians were cautious about functionality to support the completion of care and safety plans due to time pressures or perceiving these as being outside the scope of general practice. For some practices, the availability of clinicians employed under the Additional Roles Reimbursement Scheme may alleviate these tensions by increasing available human resources.

> *“More and more we try to signpost patients/customers to the right place. And [providing information about support services] is quite a useful one, ‘cause once it hits my desk there’s accountability and responsibility… and you have to dispose – using that word very loosely – of that patient. You have to. Whilst if you give them the option to look outside this GP service, first of all, so we do know what’s out there but it’s no use to us unless it can be directed to the user”* C_FG9 (clinician)

In contrast, patients/carers acknowledged the limited resources of general practice and valued it as a source of relational support and facilitating referrals, rather than to provide specialised or ongoing care. They recognised that expectations of general practice vary between patients and may initially be high, but framed expectation-setting as a two-way process enabled by compassionate, open communication without a need to limit service offerings.

> *“Maybe some [guidance around expectations] would be useful in the fact that this isn’t about waving a magic wand and self-harm will disappear because that just won’t happen. So, how can they also help the person who has limited expectations of themselves and of the services… In some cases what might be more useful is what they’re asking the person, how they are engaging in that conversation with a patient, not necessarily coming up with solutions because that could make a massive difference in somebody’s life instead of, oh they gave me the leaffet or they’ve sent me back, they’ve asked me about the thing that I already know.”* PC_FG8 (patient/carer)

## Discussion

### Summary

This study has discerned clinicians’, patients’ and carers’ needs for the content and functionality of a novel digital knowledge support tool to inform general practice care for self-harm. The themes highlight the potential for a knowledge support tool to address unmet needs across both the management and assessment of self-harm. For patients/carers, requirements of the tool were centred on building clinician-patient relationships and promoting patient choice and control. Whilst clinicians shared these priorities, they emphasised that they must be balanced with managing patient expectations and workload, and the need to practice defensively under limited resources. Some tensions were present across clinicians regarding the incorporation of risk tools.

As user requirements, these findings should inform the core design of the knowledge support tool for self-harm, which may include: (1) a purpose-built summary of patients’ health records, (2) communication prompts to support compassionate and holistic care and promote patient choice and control, (3) a tailored repository of support options, and (4) accessible leaflets to support patients and/or carers outside of consultations. Central to this, the tool’s functionality must integrate with existing workflows and allow for flexible use, and its content must be co-designed with people with lived experience of self-harm.

### Strengths and limitations

This study gave key stakeholders a decision-making role in the early design process of the knowledge support tool, thereby increasing its potential to achieve real-world impact on health outcomes.(19)

Key strengths of the study were involvement throughout of people with lived experience (including as co-authors) and a mixed-methods approach, combining qualitative and quantitative data from rating scales, focus groups and follow-on interviews with a high retention rate. This approach supports robust and rich insights through enabling data triangulation, researcher-participant rapport, reflexivity and by allowing multiple perspective to inform interpretation of findings. The framework method enabled systematic comparison of views within and across clinician and patient/carer groups. The inclusivity and accessibility of our study design was strengthened through extensive lived experience involvement and collaboration with a third-sector organisation. Our research design enhanced rigor, robustness and transparency for qualitative research (e.g. credibility, transferability, dependability and confirmability).(32, 33)

Staff participants represented a range of job roles and experience and reported working across areas of mixed deprivation. However, participant diversity was limited in other areas which may affect transferability of findings. Our study was limited to participants from White and Asian or Asian British ethnic groups. Additionally, patient/carer participants were predominantly female. The demographic imbalance for patients/carers may have been partly driven by recruitment being limited to the service users of the third-sector organisation. To mitigate recruitment gaps amongst patients/carers, an additional lived experience involvement meeting with targeted sampling was held (older people, people from ethnic groups other than White, and men).

### Comparison with existing literature

Given the limited research conducted on interventions to support general practice care for self-harm, this study has addressed an important evidence gap by identifying requirements of a knowledge support tool for this setting. Our findings are consistent with those from previous studies that identified unmet training and support needs among general practice clinicians around communication, assessing and managing risk, provision of support and referrals, continuity of care and implementation of best-practice guidelines.(7, 11, 18, 28, 39, 40) Patient and carer requirements were consistent with previous findings that patients perceive clinician-patient trust and patient agency as being important in mental health and self-harm services.(28, 41)

Our study found varying perceptions amongst clinicians regarding the use of risk scores in the management of self-harm, ranging from these being clinically and operationally useful to being potentially harmful. The appeal of risk scores and stratification-based triage to some clinician participants may reflect the perceived potential of these tools to simplify and standardise a complex clinical assessment in the context of limited time and resources (42) and a need to defend practice. Risk tools for suicidal behaviours are widely used,(30) contrary to national guidance that recommends against their use on the basis of poor predictive performance.(3, 10, 43, 44) Although no risk tool has been specifically developed and validated for UK general practice usage, uncertainty over the value of informing clinical decision-making using estimated risk of suicidal behaviours, together with outcome rarity, likely limits the utility of any such tool.(45)

Finally, our findings contribute to an understanding of how fair, high-quality care is conceptualised as healthcare services becoming increasingly digitised. Digital tool and artificial intelligence (AI) are now routinely embedded into general practice to support care delivery and reduce administrative pressures. However, there are known issues in how safety and equity are considered in the development and deployment of AI tools.(46–48) Much of the discourse around fairness of digital and AI tools is centred on data equity and algorithmic performance.(49–51) However, patient and carer accounts in this study highlight the additional importance of ensuring that tools support interpersonal and procedural fairness (ensuring that each person is listened to, treated compassionately and is given agency over their care). The knowledge support tool has potential to facilitate care equity through co-design with patients, carers and clinicians, incorporating requirements for compassion, patient agency and continuity (52) alongside established digital fairness metrics and criteria.(51)

### Implications for research and/or practice

These findings suggest that a knowledge support tool, designed to the requirements identified in this study, could meaningfully support general practice clinicians in the complex task of assessing and managing patients who have self-harmed. This knowledge support tool differs from other clinical decision support tools that have been developed or are in development, as it is specifically designed to manage people who have self-harmed in general practice settings, and is intended to enhance holistic, needs-based care rather than to inform specific clinical decisions or risk assessments.(53–55)

Our findings are particularly important given recent recommendations to scale up digital tools within mental health services whilst improving data access and supporting a needs-led approach.(56) The proposed knowledge support tool could contribute to this transformation by facilitating continuity, promoting patient agency, and by providing information and resources to enhance holistic, person-centred care. Through improving information accessibility and strengthening the capacity of general practice to facilitate needs-led care, the knowledge support tool may also provide opportunities for earlier intervention in the community, aligning with NHS priorities to shift care from hospital to the community and from sickness to prevention.(57)

The findings from this study will inform future refinement of a knowledge support tool prototype, co-designed with healthcare professionals and people with lived experience. Future co-design processes should be inclusive of diverse and underserved groups to ensure that the resulting prototype promotes health equity.

## Data Availability

To ensure confidentiality and reporting in-line with participant consent, additional data to those already captured in the manuscript are not available.

## Notes

## Ethics

Ethical approval was provided by the Wales Research Ethics Committee 3 (25/WA/0047). Further details can be found in Supplementary Information S5.

## Data

Illustrative quotes demonstrating how the data support the findings of this study are embedded in the main text and are included in Supplementary Table S8. To ensure confidentiality and reporting in-line with participant consent, additional data are not available.

## Acknowledgements

We are thankful to every participant who volunteered their time to share feedback on the knowledge support tool and their experiences of receiving or offering general practice services for self-harm. We also thank the third-sector organisation that supported recruitment, who we have not named to maintain participant anonymity. Finally, we thank our patient and carer advisory group (https://sites.manchester.ac.uk/ms4mh-r/) for their invaluable feedback and insights throughout the research process, as well as our clinical expert panel. Generative AI (Microsoft Copilot, GPT-5.6) was used for basic language editing, including grammar correction and suggestions on sentence restructuring to improve readability. No AI was used to generate ideas, methods, research questions, analyse study data, to generate scientific content, meaning, interpretation, or conclusions, or to write prose. The authors reviewed all AI suggestions and take full ownership of the manuscript content and its integrity.

