## Supplementary Material for "Requirements of a knowledge support tool for self-harm in general practice: a mixed-methods study with patients, carers, and clinicians"

### Supplementary Information S1: Plain English summary

**What did we aim to do, and why?**

Many people who self-harm seek help from GP services. However, patients often report poor experiences of care, and some staff have said they don't feel confident when providing care.

Giving better information to clinicians, patients and carers may help to improve GP services for people who self-harm. A 'knowledge support tool' is a piece of software on a GP practice computer. It can be used during consultations to deliver tailored information that may help support high-quality care. We do not have a knowledge support tool for self-harm, so were keen to find out from people with lived/living experience of self-harm and general practice staff about what one should look like.

**What did we do?**

In May-August 2025, we talked to 14 patients/carers and 13 clinicians to understand what they thought should be included within a knowledge support tool, how the tool should be used and if they thought the tool would be helpful.

Throughout the research, we worked closely with a large patient and public involvement group who have personal experience of self-harm and healthcare staff, and they also helped us to interpret the results.

**What did we find?**

Patients, carers and clinicians told us about three main needs for the knowledge support tool (described visually in Figure S1.1):

1) **The tool must place the patient at the centre of all decisions and conversations**. Patients, carers and clinicians told us that the tool should help GP clinicians understand patients' medical history, individual circumstances and needs. Patients and carers told us that it's important for conversations to be compassionate, and for patients to have a say in how their information is used.

2) **The tool must be easy to access and use.** Patients, carers and clinicians told us that the tool should provide clear, easy-to-read information and practical guidance to help GP clinicians provide care that meets patients' individual needs. For example, patients, carers and clinicians told us that it would be helpful if the tool provided information about services that may be available between GP appointments, and for this information to be written in a leaflet for patients and carers to refer to when needed. Clinicians told us that it's important that the tool can work alongside other tools that are already used on GP computers.

3) **The tool must support compassionate care in an environment where clinicians feel under a lot of pressure**. Clinicians said they must manage a heavy workload with little resources and were concerned that a knowledge support tool may create extra work or raise patients' expectations of GP services. However, patients and carers did not expect GP services to provide specialist or ongoing care. Instead, patients and carers felt it was important for patients to feel listened to, treated compassionately, and helped to find further support.

*Figure S1.1: What patients, carers and clinicians told us they need from a knowledge support tool*

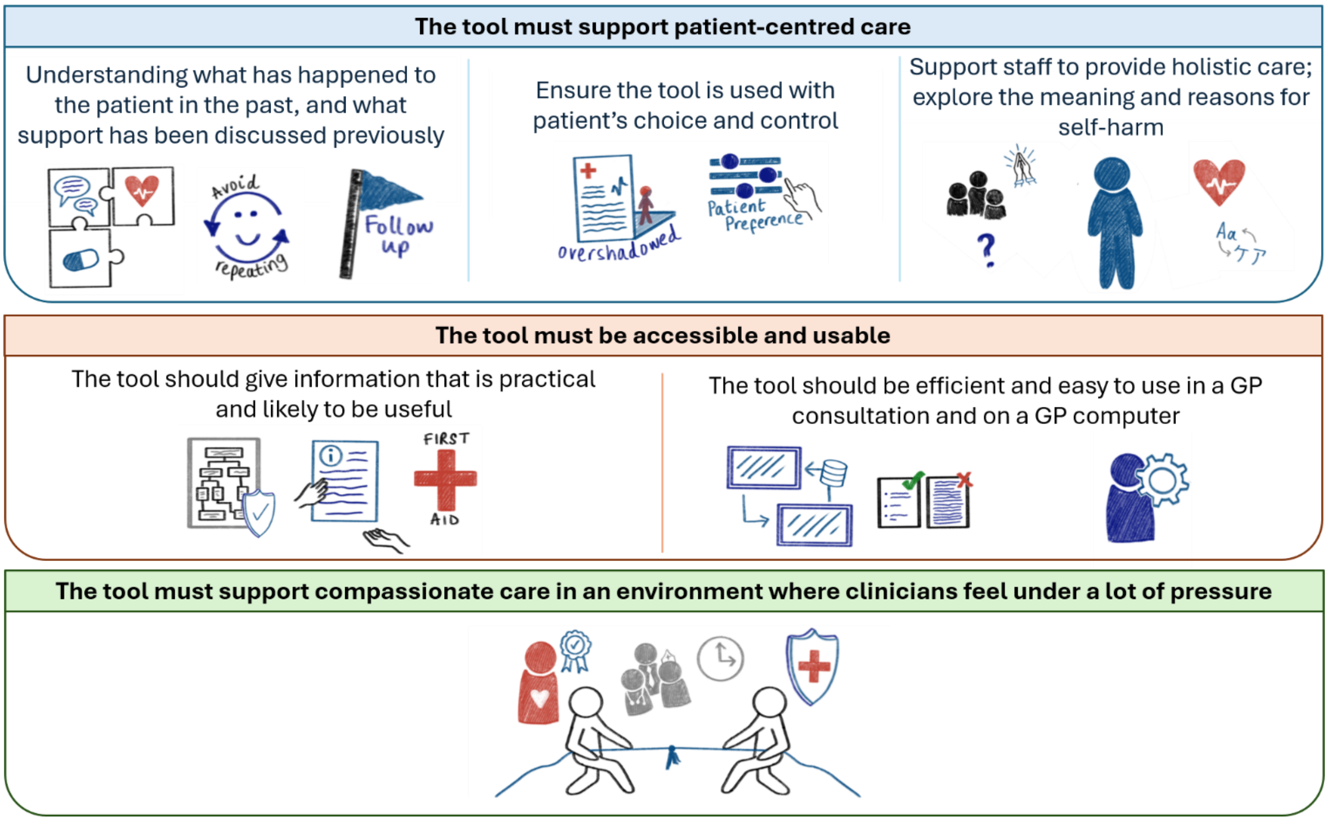


These discussions gave us an understanding of what a useful knowledge support tool for self-harm may look like. A useful knowledge support tool may include: (1) a summary of patients' health records, (2) prompts to support compassionate conversations and to help patients have more choice and control, (3) a personalised list of support options that patients may access between GP appointments, and (4) easy-to-read leaflets that patients and carers can refer to outside of appointments (e.g. containing information about different support options).

**What happens next?**

We will work with patients, carers and clinicians to develop a first prototype of the knowledge support tool. To make sure the tool is fair and helps patients equally, it is important that we speak to people from different backgrounds.

### Supplementary Information S2: Participants

Two patient/carer participants were known to some of the research team (LQ) due to prior involvement in self-harm services research. Representatives of the third-sector organisation were present during focus groups and acted as both participants and representatives in this capacity. One patient/carer participant took part in both patient/carer focus groups as they attended as both a participant and as a representative of the collaborating third-sector organisation. As recruitment of patients/carers was conducted in collaboration with the third-sector organisation, some patient/carer participants were familiar with each other and/or the representative of the third-sector organisation prior to the research study. Participants took part in a focus group and interview only once, with the exception of one patient/carer participant who was a representative of the third-sector organisation and was present during both focus groups with patients/carers.

### Supplementary Information S3: Topic guides (focus groups and interviews)

*Focus group: patient/carer*

***<u>PART 1: RATING AND DISCUSSING BROAD TYPES OF SUGGESTED CONTENT AND FUNCTIONALITY OF A KNOWELDGE SUPPORT TOOL</u>***

***1. <u>Rating exercise</u>***

***How useful would it be for a knowledge support tool to provide your doctor (or other healthcare staff) with...***

*<u>Rating guide</u>: 1=completely not useful, 5=neutral (not useful nor not useful), 10=very useful (rating scale may be visually represented for greater inclusion and understanding)*

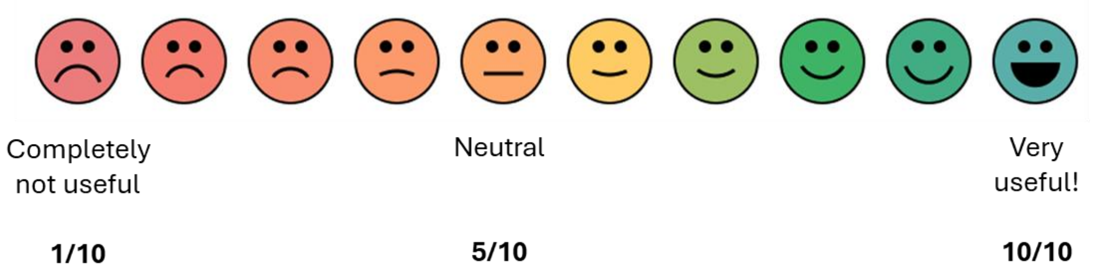


| |
|---|
| ...Information on support that may be available to you outside of GP appointments |
| ...Guidance about what to do during a GP consultation with someone who has self-harmed (e.g., based on national clinical guidance for self-harm) |
| ...Information about your health history and healthcare that you have received previously (such as hospitalisations, prescriptions and illnesses) |
| ...The ability to provide better monitoring of your health outside of GP appointments |
| ...Information leaflets about self-harm or the medicines you are taking, for you to take home after the appointment |

***2. <u>Group discussion</u>***

- Participants will be invited to join in an open discussion to talk about which content and functionality they found useful (and not useful), and why.

Facilitators will encourage participants to make suggestions to improve some of the ideas and can suggest ideas for new content and functionality altogether.

If a participant raises a new idea for content or functionality, the group will be asked for a general consensus about the usefulness of this.

***<u>PART 2: RATING AND DISCUSSING EXAMPLES OF SUGGESTED CONTENT AND FUNCTIONALITY OF A KNOWELDGE SUPPORT TOOL</u>***

***1. <u>Rating exercise</u>***

***How useful would it be for a knowledge support tool to provide your doctor (or other healthcare staff) with...***

*Rating guide: 1=completely not useful, 5=neutral (not useful nor not useful), 10=very useful (rating scale may be visually represented for greater inclusion and understanding)*

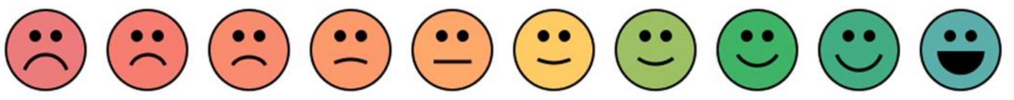

Completely not useful | Neutral | Very useful!

**1/10** | **5/10** | **10/10**

| Type of content/functionality | Examples |
|---|---|
| Information on support that may be available to you outside of GP appointments | ...Information about NHS referral pathways |
| | ... Information about community or third-sector support (regional or national) |
| | ... Information about helplines |
| Guidance about what to do during a consultation with someone who has self-harmed (e.g., based on national clinical guidance, NICE guidelines for self-harm) | ...Prompts to help your doctor assess your risk of self-harm or suicide |
| | ...Prompts for your doctor to discuss your individual care needs (including care plans) |
| | ...Guidance about what treatment or assessments you should be receiving |
| Information about healthcare that you have received previously (such as hospitalisations, prescriptions and illnesses) | ...A summary of what medicines you are currently prescribed, or used to be prescribed |
| | ...A summary of what conditions you have been diagnosed with |
| | ...A summary of services you have been previously or currently accessed (including hospital visits and referrals) |
| The ability to provide better monitoring of your health outside of GP appointments | ...Surveys or apps that you can use to log your mood, thoughts related to self-harm and suicidal ideation |
| Information leaflets about self-harm or the medicines you are taking, for you to take home after the appointment | ...Information leaflets that explain what self-harm is or give advice on how to manage self-harm, for you to take home after the appointment |
| | ...Information leaflets that explain what medicines you are taking, or care that you are receiving, for you to take home after the appointment |

| | |
|---|---|
| | ...Information leaflets that give details for available support outside of GP appointments, for you to take home after the appointment |
| | ...Information leaflets in accessible/easy-read formats and different languages, for you to take home after the appointment |

**2. *Group discussion***

- Similar format as for the first group discussion

*Focus group: general practice clinicians*

***PART 1: RATING AND DISCUSSING BROAD TYPES OF SUGGESTED CONTENT AND FUNCTIONALITY OF A KNOWELDGE SUPPORT TOOL***

**1. *Rating exercise***

***How useful would it be for a knowledge support tool to provide healthcare staff with...***

*Rating guide: 1=completely not useful, 5=neutral (not useful nor not useful), 10=very useful (rating scale may be visually represented for greater inclusion and understanding)*

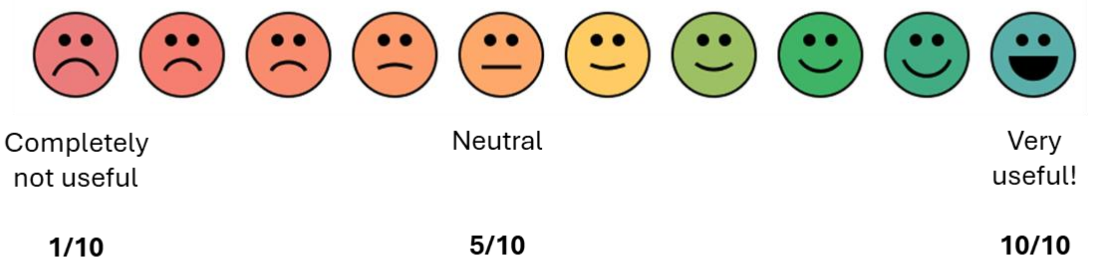


| |
|---|
| ...Information on best-practice guidelines (eg NICE) for self-harm |
| ...Prompts to support communication and documentation of self-harm |
| ...Information about a patient's medicines, care plans or clinical history |
| ...Information leaflets for patients/carers to refer to after leaving the GP practice |
| ...Information on available support for patients outside of GP appointments |
| ...The ability to provide better monitoring of patients outside of GP appointments |

**2. *Group discussion***

- Participants will be invited to join in an open discussion to talk about which content and functionality they found useful (and not useful), and why.

Facilitators will encourage participants to make suggestions to improve some of the ideas and can suggest ideas for new content and functionality altogether.

If a participant raises a new idea for content or functionality, the group will be asked for a general consensus about the usefulness of this.

***PART 2: RATING AND DISCUSSING EXAMPLES OF SUGGESTED CONTENT AND FUNCTIONALITY OF A KNOWELDGE SUPPORT TOOL***

***1. Rating exercise***

***How useful would it be for a knowledge support tool to provide healthcare staff with…***

*Rating guide: 1=completely not useful, 5=neutral (not useful nor not useful), 10=very useful (rating scale may be visually represented for greater inclusion and understanding)*

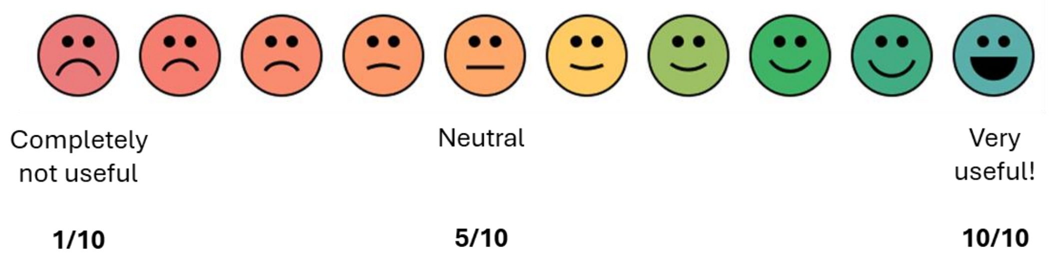


| Feature type | Examples |
|---|---|
| Information on best-practice guidelines for self-harm | …Information from NICE guidelines about self-harm management/assessment and suicide prevention |
| | …Flagging when guideline recommendations have not been met (e.g. high risk prescribing) |
| | …Information from studies/other published sources which may help guide self-harm management/assessment |
| Prompts to support communication and documentation of self-harm | …Prompts to ensure patient's self-harm is coded into their health record |
| | …Prompts to assess presence of risk factors which may be associated with harmful outcomes (such as suicide) |
| | …Prompts to discuss patient/carer individual care needs (including care plans) |
| Information about a patient's medicines, care plans or clinical history | …A summary of patients' medication prescribing history |
| | …A summary of patients' clinical events history (e.g. diagnoses) |
| | …A summary of services that have been previously or currently accessed by the patient (including hospital visits and referrals) |

| | |
|---|---|
| Information leaflets for patients/carers to refer to after leaving the GP practice | ...Information leaflets that explain what self-harm is or give advice on how to manage self-harm, for patients/carers to refer to after leaving the GP practice |
| | ...Information leaflets that explain what medicines the patient is taking, or care that they are receiving, for patients/carers to refer to after leaving the GP practice |
| | ...Information leaflets that give details for available support outside of GP appointments, for patients/carers to refer to after leaving the GP practice |
| | ...Information leaflets in accessible/easy-read formats and different languages, for patients/carers to refer to after leaving the GP practice |
| Information on available support for patients outside of GP appointments | ...Information about NHS referral pathways |
| | ...Information about community or third-sector support (regional or national) |
| | ...Information about helplines |
| The ability to provide better monitoring of patients outside of GP appointments | ...Surveys or apps that patients/carers can use to log mood, thoughts related to self-harm and suicidal ideation |

***2. <u>Group discussion</u>***

- Similar format as for the first group discussion

*Interview: patient/carer*

***<u>PART 1: REQUIREMENTS FOR CONTENT AND FUNCTIONALITY OF A KNOWELDGE SUPPORT TOOL</u>***

- Facilitators will present a summary of the requirements and perspectives on content and functionality that were identified during the focus groups. The participant will be given the opportunity to ask any questions and provide further individual views on the features of a knowledge support tool for self-harm.
- This will take the format of an open discussion. If the participant does not have any comments to add, that is okay.

***<u>PART 2: THEORETICAL FRAMEWORK OF ACCEPTABILITY</u>***

- Each question will invite the participant to rate how they feel about a different aspect of acceptability of the knowledge support system. After giving a rating, participants will be invited to engage in a discussion about their views and feelings, to comment on or contextualise their rating. Participants may at any point ask for further clarification about what a question is asking.

*<u>Rating guide</u>: All ratings are given on a 10-point scale (rating scale may be visually represented for greater inclusion and understanding)*

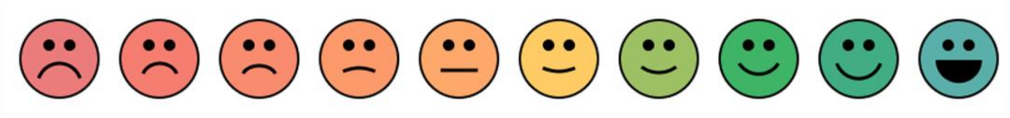

**1/10** **5/10** **10/10**

1. *<u>'Affective attitude' (feelings towards the tool):</u>*
   How comfortable would you feel if a knowledge support tool was used during your GP appointment relating to self-harm?

| Very uncomfortable | Neutral | Very comfortable |
|---|---|---|
| 1 | 5 | 10 |

2. *<u>'Burden' (how hard or tiring something is):</u>*
   How much effort do you think it will take to use the knowledge support tool during a GP appointment relating to self-harm?

| No effort at all | Neutral | Huge effort |
|---|---|---|
| 1 | 5 | 10 |

3. *<u>'Ethicality' (e.g., if something is done in a fair, safe, respectful, and fits with the how you think, feel, and act):</u>*
   Do you think using a knowledge support tool to inform GP appointments for self-harm is a good fit for your personal values (how you think, feel and act)?

| Very bad fit | Neutral | Very good fit |
|---|---|---|
| 1 | 5 | 10 |

4. *<u>'Perceived effectiveness'(how much you think/believe the tool is working):</u>*
   How confident are you that a knowledge support tool will provide the knowledge needed to improve the care of people who have self-harmed in GP practices?

| Very unconfident | Neutral | Very confident |
|---|---|---|
| 1 | 5 | 10 |

5. *<u>'Intervention coherence' (does it make sense to you)</u>*:
   How clear is it to you *how* the knowledge support tool might help inform healthcare staff and patients or carers?

| Very unclear | Neutral | Very clear |
|---|---|---|
| 1 | 5 | 10 |

6. *<u>'Self-efficacy' (belief in the GP that they can use the tool)</u>*:
   How confident are you that healthcare staff, patients and carers will be able to use the knowledge support tool?

| Very unconfident | Neutral | Very confident |
|---|---|---|
| 1 | 5 | 10 |

7. *<u>'Opportunity costs' (what is given up to do the tool, e.g., time/money)</u>*:
   Do you think that the knowledge support tool being used during an appointment relating to self-harm would interfere with other things are important to you?

| No extent | Neutral | Huge extent |
|---|---|---|
| 1 | 5 | 10 |

8. *<u>'Safety':</u>*
   Do you think using the knowledge support tool to inform GP appointments relating to self-harm is safe?

| Very unsafe | Neutral | Very safe |
|---|---|---|
| 1 | 5 | 10 |

9. *<u>'Overall acceptability' (your overall opinion of the tool, considering everything we have spoken about, e.g., easy, comfortable, works well, like/dislike):</u>*
   Overall, do you think that using a knowledge support tool to help inform GP appointments relating to self-harm is acceptable?

| Completely unacceptable | Neutral | Completely acceptable |
|---|---|---|
| 1 | 5 | 10 |

***<u>PART 3: IMPLEMENTATION OF A KNOWLEDGE SUPPORT TOOL</u>***

1. Do you have any preferences on *when* the knowledge support tool should be used?
2. Do you have any preferences on *who* should use the knowledge support tool in general practice?
3. Do you have any preferences on who the knowledge support tool could be used *for*?
4. What do you think might prevent patients and carers approving the use of this knowledge support tool? What might help address these concerns?
5. What do you think might prevent healthcare staff from using the knowledge support tool? What might help address these concerns?

*Interview: general practice clincians*

***PART 1: REQUIREMENTS FOR CONTENT AND FUNCTIONALITY OF A KNOWELDGE SUPPORT TOOL***

- Facilitators will present a summary of the requirements and perspectives on content and functionality that were identified during the focus groups. The participant will be given the opportunity to ask any questions and provide further individual views on the features of a knowledge support tool for self-harm.
- This will take the format of an open discussion. If the participant does not have any comments to add, that is okay.

***PART 2: THEORETICAL FRAMEWORK OF ACCEPTABILITY***

- Each question will invite the participant to rate how they feel about a different aspect of acceptability of the knowledge support system. After giving a rating, participants will be invited to engage in a discussion about their views and feelings, to comment on or contextualise their rating. Participants may at any point ask for further clarification about what a question is asking.

*Rating guide: All ratings are given on a 10-point scale (rating scale may be visually represented for greater inclusion and understanding)*

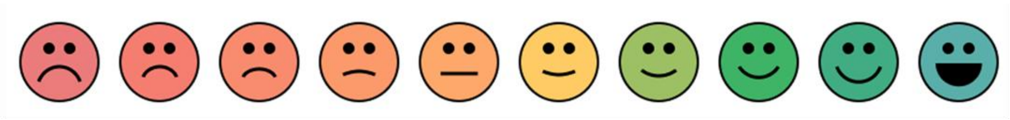

**1/10** **5/10** **10/10**

1. *‘Affective attitude’ (feelings towards the tool):*
   How comfortable would you using a knowledge support tool during an appointment relating to self-harm?

| Very uncomfortable | Neutral | Very comfortable |
|---|---|---|
| 1 | 5 | 10 |

2. *‘Burden’ (how hard or tiring something is):*
   How much effort do you think it will take to use the knowledge support tool during an appointment relating to self-harm?

| No effort at all | Neutral | Huge effort |
|---|---|---|
| 1 | 5 | 10 |

3. *‘Ethicality’ (e.g., if something is done in a fair, safe, respectful, and fits with the how you think, feel, and act):*

Do you think using a knowledge support tool to inform consultations for self-harm is a good fit for your personal values (how you think, feel and act)?

| Very bad fit | Neutral | Very good fit |
|---|---|---|
| 1 | 5 | 10 |

*4.* *‘Perceived effectiveness’(how much you think/believe the tool is working):*
How confident are you that a knowledge support tool will provide the knowledge needed to improve the care of people who have self-harmed in GP practices?

| Very unconfident | Neutral | Very confident |
|---|---|---|
| 1 | 5 | 10 |

5. *‘Intervention coherence’ (does it make sense to you)*:
How clear is it to you how the knowledge support tool might help inform healthcare staff and patients or carers?

| Very unclear | Neutral | Very clear |
|---|---|---|
| 1 | 5 | 10 |

6. *‘Self-efficacy’ (belief in the GP that they can use the tool)*:
How confident are you that healthcare staff, patients and carers will be able to use the knowledge support tool?

| Very unconfident | Neutral | Very confident |
|---|---|---|
| 1 | 5 | 10 |

*7.* *‘Opportunity costs’ (what is given up to do the tool, e.g., time/money)*:
To what extent would using the knowledge support tool during an appointment relating to self-harm interfere with other things are important to you?

| No extent | Neutral | Huge extent |
|---|---|---|
| 1 | 5 | 10 |

*8.* *‘Safety’:*
Do you think using the knowledge support tool to inform GP appointments relating to self-harm is safe?

| Very unsafe | Neutral | Very safe |
|---|---|---|
| 1 | 5 | 10 |

*9.* *‘Overall acceptability’ (your overall opinion of the tool, considering everything we have spoken about, e.g., easy, comfortable, works well, like/dislike):*
Overall, do you think that using a knowledge support tool to help inform GP appointments relating to self-harm is acceptable?

| Completely unacceptable | Neutral | Completely acceptable |
| --- | --- | --- |
| 1 | 5 | 10 |

***PART 3: IMPLEMENTATION OF A KNOWLEDGE SUPPORT TOOL***

1. Do you have any preferences on *when* the knowledge support tool should be used?
2. Do you have any preferences on *who* should use the knowledge support tool in general practice?
3. Do you have any preferences on who the knowledge support tool could be used *for*?
4. What do you believe the incentives/disincentives of using the knowledge support tool are, and do you think that these are sufficient to encourage the use of the knowledge support tool?
5. What do you think might prevent patients and carers approving the use of this knowledge support tool? What might help address these concerns?
6. What resources additional to what you have now are needed to be able to use the knowledge support tool? What other resources might you need?
7. Finally, do you have any other comments on what you think might prevent successful and sustainable implementation of the knowledge support tool in practice?

### Supplementary Information S4: Inclusion, accessibility and participant safety and wellbeing

Our approach to recruitment and data collection was centred around disability and neurodivergent inclusivity.(1-3) All topic guide questions were asked both orally and as written questions and were accompanied by visual anchors (Supplementary Information S3). Participants were given agency to contribute orally or through written responses. For patient/carer participants, in-person data collection was offered at a familiar and accessible community location where sensory fidgets and art-based activities were available. A representative from the third-sector organisation was present during lived experience focus groups to support participants' wellbeing and accessibility if needed, for example by anticipating and supporting individual communication needs. A representative from the third sector was also available at the participants' request for the follow-on interviews (no participant requested this).

A risk protocol was developed that detailed the approach to be taken if a participant showed signs of distress. For patient/carers, the study was carried out in collaboration with a third-sector organisation that provide support services for people who have been affected by self-harm. All participants were given details for additional support services prior to the study, which were tailored separately for clinician and patient/carer participants.

Prior to publication, consenting participants were sent a summary of the study findings.

1. McLeod J. Why we interview now--reflexivity and perspective in a longitudinal study. International Journal of Social Research Methodology. 2003;6(3):201-11.
2. Calman L, Brunton L, Molassiotis A. Developing longitudinal qualitative designs: lessons learned and recommendations for health services research. BMC Medical Research Methodology. 2013;13(1):14.
3. Lloyd N, Hyett N, Kenny A. To Member Check or not to Member Check? An Evaluation of Member Checking in an Interpretive Descriptive Study. International Journal of Qualitative Methods. 2024;23:16094069241301383.

Supplementary Material Figures S1-S2: Visual focus group summaries

*Figure S1: Visual summary of patient/carer focus group discussions*

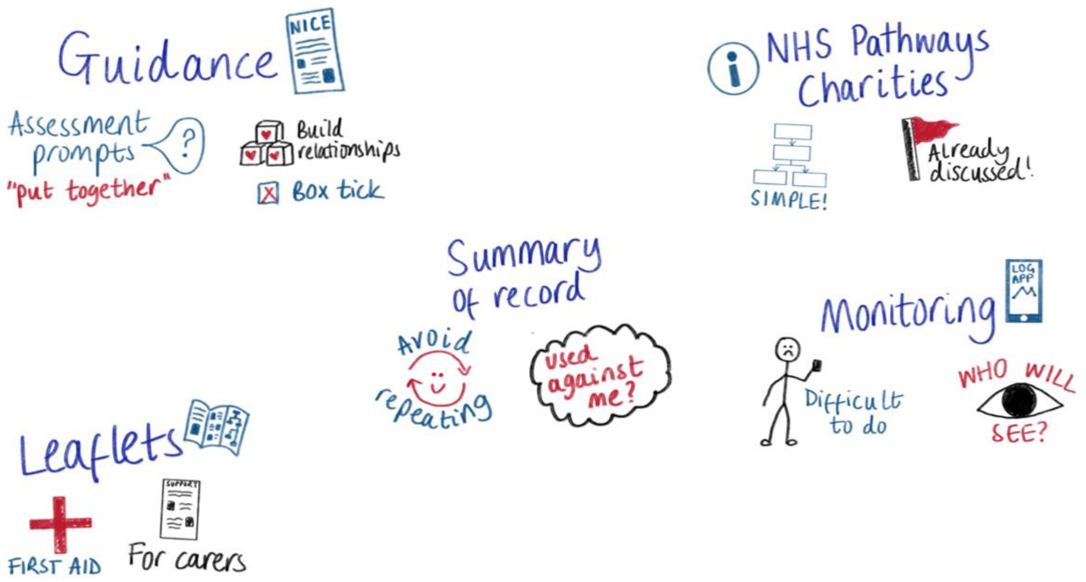


*Figure S2: Visual summary of general practice clinician focus group discussions*

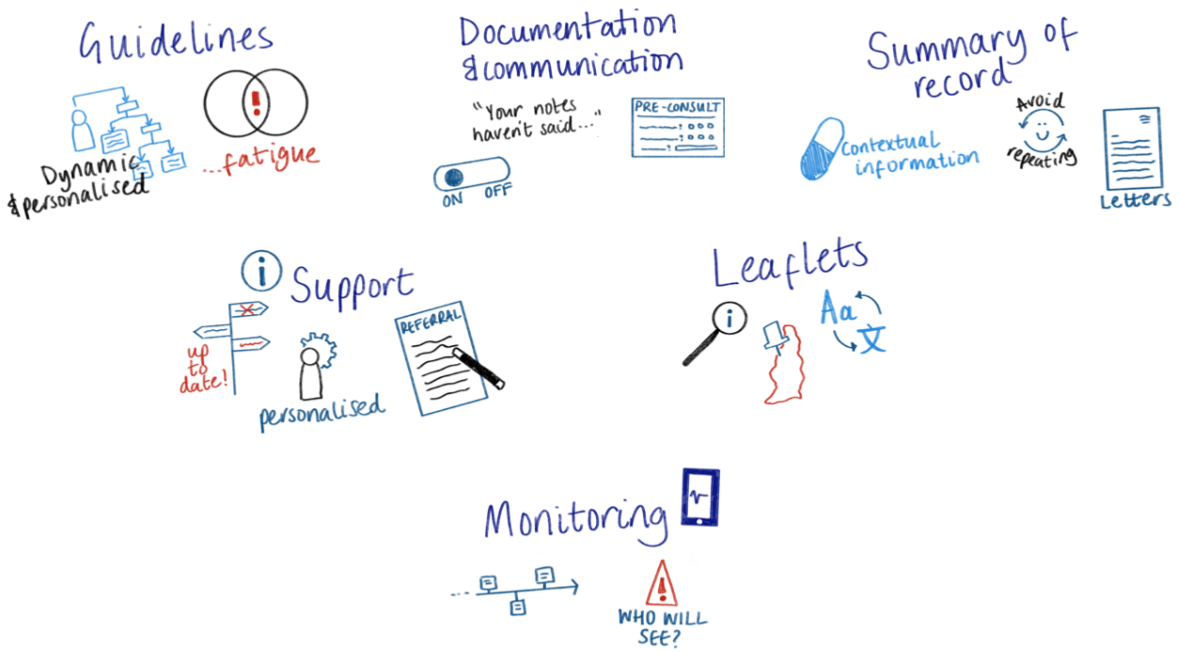

### Supplementary Table S1: Criteria for trustworthiness

*Table S1: Approaches taken to fulfil criteria for trustworthiness (Lincoln and Guba)*

| **Approach** | **Criteria** |
|---|---|
| The longitudinal study design (focus groups followed by individual interviews) allowed for both participant reflexivity (i.e. allowing participants to reflect on changing viewpoints after a period of reflection and familiarity of the KS tool) and member checking (confirming researcher interpretation of focus group findings during the interviews).(1-3) It also allowed a rapport between facilitators and participants to develop prior to individual interviews, which may facilitate deeper insights.(4) | Credibility, confirmability |
| Investigator triangulation was employed throughout the analysis lifecycle. Findings were shared and discussed within multidisciplinary teams consisting of researchers with expertise in digital tool development (HC, TvS), health services research (HC, LQ, TvS, NK, FM, RW, JW), self-harm research (LQ, NK, FM, RW, JW), qualitative research (LQ, FM; HC additionally has attended multiple training modules on qualitative methods), a lived-experience co-applicant and co-authors (SA, JG, EG) and a panel of healthcare professional and lived experience contributors (including SA and KP). An additional lived experience involvement meeting was held with targeted sampling to mitigate recruitment gaps (older people, people from ethnic minority backgrounds, men). Investigator triangulation ensured that data interpretations considered clinical and lived experience perspectives, and validated findings in broader populations.(5) | Credibility, transferability, confirmability |
| Rich description was facilitated through use of framework matrices, which involved charting participant data against developed codes/themes through use of quotations and summaries, and additionally allowed identification of outlier cases.(6) Rich description was also facilitated through in-depth description of participant demographics, allowing for interpretation of whether findings might be applicable to other populations.(4) | Credibility, transferability |

| | |
|---|---|
| Prior to and following each focus group and interview, facilitators (who also led data analysis) HC and LQ met to reflexively reflect on discussions, including how their own positionalities may affect interpretation.(5) | Credibility |
| A clear audit trail of the analysis steps was produced to provide a reference log of decisions and changes in coding approaches and frameworks.(4) Additionally, topic guides have been published for transparency. | Dependability, transferability, confirmability |

Based on Lincoln and Guba's criteria for trustworthiness (7)

1. McLeod J. Why we interview now--reflexivity and perspective in a longitudinal study. International Journal of Social Research Methodology. 2003;6(3):201-11.
2. Calman L, Brunton L, Molassiotis A. Developing longitudinal qualitative designs: lessons learned and recommendations for health services research. BMC Medical Research Methodology. 2013;13(1):14.
3. Lloyd N, Hyett N, Kenny A. To Member Check or not to Member Check? An Evaluation of Member Checking in an Interpretive Descriptive Study. International Journal of Qualitative Methods. 2024;23:16094069241301383.
4. Ahmed SK. The pillars of trustworthiness in qualitative research. J Med Surg Public Health. 2024;2:100051.
5. Rose J, Johnson CW. Contextualizing reliability and validity in qualitative research: toward more rigorous and trustworthy qualitative social science in leisure research. Journal of Leisure Research. 2020;51(4):432-51.
6. Gale NK, Heath G, Cameron E, Rashid S, Redwood S. Using the framework method for the analysis of qualitative data in multi-disciplinary health research. BMC Med Res Methodol. 2013;13:117.
7. Lincoln YS, Guba EG. Naturalistic inquiry. SAGE Publications. 1985.

### Supplementary Table S2: Coding framework

*Table S2: Coding framework informing the qualitative themes*

| | |
|---|---|
| Patient record | |
| | Accuracy of record |
| | Bias |
| | Care plans |
| | Diagnoses |
| | Existing clinician access |
| | Extent and method of summarising |
| | Integrated records |
| | Letters and free text |
| | Medicolegal |
| | Other clinical events |
| | Overshadowing |
| | Prescribing |
| | Services accessed |
| | Use varying by clinician |
| | Patient record benefit |
| Monitoring | |
| | Actions in response |
| | Clinical responsibility and medicolegal |
| | Input prior to consultation |
| | Monitoring benefit |
| | Negative emotional response |
| | Practicality |
| | Trust |
| Guidance and prompts | |

| |
|---|
| Appropriate language |
| Carers |
| Liability and medicolegal |
| NICE guidance |
| Guideline deviations |
| Guidance outside of NICE |
| Treatments and care plans |
| Patient-centred care |
| Practicality and format |
| Relational |
| Guidance and prompts benefit |
| Risk and assessments |
| External support |
| Better information and language |
| Carers |
| Continuity |
| Helplines |
| Individualised support |
| NHS referral pathways |
| Third sector support |
| Practicality |
| External support benefit |
| Leaflets |
| Carers |
| General self harm |
| Harm minimisation |
| Patient-centred care |
| Patient's care |
| Support |

| |
|---|
| Ensuring relevance |
| Format and language |
| Existing resources |
| Leaflets benefit |

### Supplementary Table S3: Illustration of how quantitative results aligned with and informed qualitative themes

*Table S3: Illustration of how quantitative results aligned with and informed qualitative results*

| Theme/subtheme | Qualitative finding | Informative utility ratings for suggested content and functionality of a knowledge support tool (types and examples), presented using median (interquartile range) |
|---|---|---|
| Theme 1: Content to facilitate person-centred care | | |
| Subtheme 1: Building informational and management continuity | "Both clinicians and patients/carers valued summaries of hard-to-access information from the patients' health record" | Information about a patient's medicines, care plans or clinical history:<br>- Staff: 9.0 (5.0-10.0)<br>- Patients/carers: 6.0 (5.0-10.0)<br>A summary of clinical events history (e.g. diagnoses):<br>- Staff: 10.0 (7.0-10.0)<br>- Patients/carers*: 5.5 (4.2-9.8)<br>A summary of medication prescribing history:<br>- Staff: 10.0 (8.5-10.0)<br>- Patients/carers: 8.0 (7.5-10.0)<br>A summary of services that have been previously or currently accessed (including hospital visits and referrals):<br>- Staff: 10.0 (8.0-10.0)<br>- Patients/carers: 8.0 (6.0-9.5) |
| | "Some patients/carers additionally suggested that the tool may support management continuity by recording which support options have been previously discussed, prompting clinicians to follow-up with ongoing care and avoid offering | Information on helplines:<br>- Patients/carers: 6.0 (5.0-8.5) |

| | | |
|---|---|---|
| | management options that patients are already aware of or had identified as unhelpful” | |
| Subtheme 2: Responsive to patients’ choice and control | “Whilst patients/carers valued summaries of patients’ records, they expressed concern that summarising historical diagnoses may cause clinicians to make assumptions about the cause of their self-harm or enable potential misdiagnoses to influence care.” | A summary of clinical events history (e.g. diagnoses)*:<br>- Patients/carers: 5.5 (4.2-9.8) |
| | “Similarly, patients/carers were concerned that functionality enabling patients to log their mood and thoughts outside consultations could reduce patients’ control over who can access or use this information” | The ability to provide better monitoring of your health outside of GP appointments:<br>- Patients/carers: 5.5 (4.0-6.8)<br>Surveys or apps that you can use to log your mood, thoughts related to self-harm and suicidal ideation:<br>- Patients/carers: 3.0 (1.2-4.5) |
| Subtheme 3: Supporting holistic care: opportunities and tensions | “Both participant groups emphasised the importance of adapting the knowledge support tool content to meet patients’ cultural and language needs, for example through translated patient leaflets and prompts to consider religion”<br>Information leaflets in accessible/easy-read formats and different languages: | - Staff: 10.0 (10.0-10.0)<br>- Patients/carers: 10.0 (9.0-10.0) |
| Theme 2: Accessibility and usability | | |
| Subtheme 1: Practical and actionable | “Staff and patients/carers valued content to promote care aligned with up-to-date guidance from the National Institute for Health and Care Excellence (NICE)” | Information on best-practice guidelines (eg NICE) for self-harm:<br>- Staff: 7.0 (7.0-9.0)<br>Information from NICE guidelines about self-harm management/assessment and suicide prevention: |

| | | |
|---|---|---|
| | | - Staff: 7.0 (5.0-8.0)<br>Flagging when guideline recommendations have not been met (e.g. high risk prescribing):<br>- Staff: 8.0 (7.0-10.0)<br>Guidance about what to do during a GP consultation with someone who has self-harmed (e.g., based on national clinical guidance for self-harm):<br>- Patients/carers: 10.0 (8.0-10.0) |
| | "For example, participants from both groups welcomed information on available support services, and for this information to be detailed in leaflets that patients and carers may refer to when needed." | Information leaflets that give details for available support outside of GP appointments:<br>- Staff: 10.0 (9.0-10.0)<br>- Patients/carers: 8.0 (7.5-10.0)<br>Information leaflets that explain what medicines the patient is taking, or care that they are receiving:<br>- Staff: 9.0 (8.5-10.0)<br>Patients/carers: 8.0 (6.5-8.5) |
| | "Both clinicians and patients/carers also viewed communication prompts (e.g. suggested discussion points or guidance on phrasing or language) as useful to directly support conversations around care needs and risk factors" | Prompts to discuss patient/carer individual care needs (including care plans):<br>- Staff: 8.0 (6.0-9.0)<br>- Patients/carers: 8.0 (6.5-9.0) |
| Subtheme 2: Streamlining workflows | "Staff valued functionality to automate time-consuming administrative processes, such as aiding documentation of consultations and collating up-to-date details for support services." | Prompts to support communication and documentation of self-harm:<br>- Staff: 9.0 (5.0-10.0)<br>Prompts to ensure patient's self-harm is coded into their health record:<br>Staff: 8.0 (5.0-9.0) |
| Theme 3: Balancing competing priorities of general practice | | |

| | | |
|---|---|---|
| Theme 3: Balancing competing priorities of general practice | "As such, clinicians expressed concerns about functionality that allows patients to self-report symptoms or distress between consultations, as this may place additional expectation on general practice to respond to them in real-time or carry medicolegal implications if they do not" | The ability to provide better monitoring of patients outside of GP appointments:<br>- Staff: 5.0 (5.0-6.0)<br>Surveys or apps that patients/carers can use to log mood, thoughts related to self-harm and suicidal ideation:<br>- Staff: 5.0 (2.5-5.0) |

* Patients/carers were instead presented with 'summary of conditions a patient has been diagnosed with'; this narrower scope was intended to reduce conceptual breadth and increase clarity.

### Supplementary Tables S4-6: Reporting standards

#### *SRQR*

*Table S4: Demonstration of meeting reporting criteria: Standards for Reporting Qualitative Research*

| **No. Topic** | **Item** | **Page number(s) – submitted manuscript** |
|---|---|---|
| **Title and abstract** | | |
| S1 Title | Concise description of the nature and topic of the study identifying the study as qualitative or indicating the approach (e.g., ethnography, grounded theory) or data collection methods (e.g., interview, focus group) is recommended | 1 |
| S2 Abstract | Summary of key elements of the study using the abstract format of the intended publication; typically includes objective, methods, results, and conclusions | 2 |
| **Introduction** | | |
| S3 Problem formulation | Description and significance of the problem/phenomenon studied; review of relevant theory and empirical work; problem statement | 5-6 |

| | | |
|---|---|---|
| S4 Purpose or research question | Purpose of the study and specific objectives or questions | 6 |
| **Methods** | | |
| S5 Qualitative approach and research paradigm | Qualitative approach (e.g., ethnography, grounded theory, case study, phenomenology, narrative research) and guiding theory if appropriate; identifying the research paradigm (e.g., positivist, constructivist/interpretivist) is also recommended | 7 |
| S6 Researcher characteristics and reflexivity | Researchers' characteristics that may influence the research, including personal attributes, qualifications/experience, relationship with participants, assumptions, or presuppositions; potential or actual interaction between researchers' characteristics and the research questions, approach, methods, results, or transferability | 8;<br>Supplementary Information S2 |
| S7 Context | Setting/site and salient contextual factors; rationale | 7-8 |
| S8 Sampling strategy | How and why research participants, documents, or events were selected; criteria for deciding when no further sampling was necessary (e.g., sampling saturation); rationale | 7 |
| S9 Ethical issues pertaining to human subjects | Documentation of approval by an appropriate ethics review board and participant consent, or explanation for lack thereof; other confidentiality and data security issues | 23;<br>Supplementary Information S5 |

| | | |
|---|---|---|
| S10 Data collection methods | Types of data collected; details of data collection procedures including (as appropriate) start and stop dates of data collection and analysis, iterative process, triangulation of sources/methods, and modification of procedures in response to evolving study findings; rationale | 167-236;<br>Supplementary Table S1 |
| S11 Data collection instruments and technologies | Description of instruments (e.g., interview guides, questionnaires) and devices (e.g., audio recorders) used for data collection; if/how the instrument(s) changed over the course of the study | 8-9;<br>Supplementary Information S3 |
| S12 Units of study | Number and relevant characteristics of participants, documents, or events included in the study; level of participation (could be reported in results) | 11-12;<br>Supplementary Figure S3 |
| S13 Data processing | Methods for processing data prior to and during analysis, including transcription, data entry, data management and security, verification of data integrity, data coding, and anonymization/deidentification of excerpts | 9-10;<br>Supplementary Information S5 |
| S14 Data analysis | Process by which inferences, themes, etc., were identified and developed, including researchers involved in data analysis; usually references a specific paradigm or approach; rationale | 7;<br>9-10 |
| S15 Techniques to enhance trustworthiness | Techniques to enhance trustworthiness and credibility of data analysis (e.g., member checking, audit trail, triangulation); rationale | Supplementary Table S1 |
| **Results/Findings** | | |

| | | |
|---|---|---|
| S16 Synthesis and interpretation | Main findings (e.g., interpretations, inferences, and themes); might include development of a theory or model, or integration with prior research or theory | 11-19; |
| S17 Links to empirical data | Evidence (e.g., quotes, field notes, text excerpts, photographs) to substantiate analytic findings | 11-19;<br>Supplementary Tables S3, S7, S8 |
| **Discussion** | | |
| S18 Integration with prior work, implications, transferability, and contribution(s) to the field | Short summary of main findings; explanation of how findings and conclusions connect to, support, elaborate on, or challenge conclusions of earlier scholarship; discussion of scope of application/generalizability; identification of unique contribution(s) to scholarship in a discipline or field | 20-22 |
| S19 Limitations | Trustworthiness and limitations of findings | 20-21;<br>Supplementary Table S1 |
| **Other** | | |
| S20 Conflicts of interest | Potential sources of influence or perceived influence on study conduct and conclusions; how these were managed | 23 |
| S21 Funding | Sources of funding and other support; role of funders in data collection, interpretation, and reporting | 23 |

Standards for Reporting Qualitative Research (SRQR): O'Brien BC, Harris IB, Beckman TJ, Reed DA, Cook DA. Standards for Reporting Qualitative Research: A Synthesis of Recommendations. Acad Med. 2014;89(9):1245-51.

#### *JARS-Mixed*

*Table S5: Demonstration of meeting reporting criteria: Journal Article Reporting Standards for mixed-methods research (JARS-Mixed)*

| **Topic** | **Page number – submitted manuscript** |
|---|---|
| *Abstract* | |
| Indicate the mixed methods design, including types of participants or data sources, analytic strategy, main results/findings, and major implications/significance. | 2 |
| *Introduction* | |
| State three types of research objectives/aims/goals: qualitative, quantitative, and mixed methods. Order these goals to reflect the type of mixed methods design used. | N/A – all are related to the stated research question |
| Describe the ways approaches to inquiry were combined, as it illuminates the objectives and mixed methods rationale (e.g., descriptive, interpretive, feminist, psychoanalytic, postpositivist, critical, postmodern, constructivist, or pragmatic approaches). | 7 |
| *Method* | |
| Explain why mixed methods research is appropriate as a methodology given the paper's goals. | 7 |
| Identify the type of mixed methods design used and define it. | 7 |
| Indicate the qualitative approach to inquiry and the quantitative design used within the mixed methods design type (e.g., ethnography, randomized experiment). | 7 |

| | |
|---|---|
| If multiple approaches to inquiry were combined, describe how this was done and provide a rationale (e.g., descriptive, interpretive, feminist, psychoanalytic, postpositivist, critical, postmodern, constructivist, or pragmatic approaches), as it is illuminating for the mixed method in use. | N/A |
| Provide a rationale or justification for the need to collect both qualitative and quantitative data and the added value of integrating the results (findings) from the two databases. | 7 |
| When data are collected from multiple sources, clearly identify the sources of qualitative and quantitative data (e.g., participants, text), their characteristics, and the relationship between the data sets, if there is one (e.g., an embedded design). | 8-9 |
| State the data sources in the order of procedures used in the design type (e.g., qualitative sources first in an exploratory sequential design followed by quantitative sources), if a sequenced design is used in the mixed methods study. | 7-9 |
| Describe the qualitative and the quantitative sampling in separate sections. | N/A – not conducted separately |
| Relate the order of the sections to the procedures used in the mixed methods design type. | 7-9 |
| Discuss the recruitment strategy for qualitative and quantitative research separately | N/A – not conducted separately |
| Devote separate sections to the qualitative data analysis, the quantitative data analysis, and the mixed methods analysis. This mixed methods analysis consists of ways that the quantitative and qualitative results will be “mixed” or integrated according to the type of mixed methods design used (e.g., merged in a convergent design, connected in explanatory sequential designs and in exploratory sequential designs). | 8-9 |

| | |
|---|---|
| Indicate methodological integrity, quantitative validity and reliability, and mixed methods validity or legitimacy. Further assessments of mixed methods integrity are also indicated to show the quality of the research process and the inferences drawn from the intersection of the quantitative and qualitative data. | Supplementary Table S2-S3 |
| *Findings/results* | |
| Indicate how the qualitative and quantitative results were "mixed" or integrated (e.g., discussion; tables of joint displays; graphs; data transformation in which one form of data is transformed to the other, such as qualitative text, codes, themes are transformed into quantitative counts or variables). | 9-10;<br>Supplementary Table S3 |

Journal Article Reporting Standards for mixed-methods research (JARS-Mixed): Levitt HM, Bamberg M, Creswell JW, Frost DM, Josselson R, Suárez-Orozco C. Journal article reporting standards for qualitative primary, qualitative meta-analytic, and mixed methods research in psychology: The APA Publications and Communications Board task force report. Am Psychol. 2018;73(1):26-46.

### *COREQ*

*Table S6: Demonstration of meeting reporting criteria: Consolidated criteria for reporting qualitative research (COREQ)*

| Topic | Item No. | Guide Questions/Description | Page number – submitted manuscript |
|---|---|---|---|
| **Domain 1: Research team and reflexivity** | | | |
| *Personal characteristics* | | | |
| Interviewer/facilitator | 1 | Which author/s conducted the interview or focus group? | 8 |
| Credentials | 2 | What were the researcher's credentials? E.g. PhD, MD | 1-2 |
| Occupation | 3 | What was their occupation at the time of the study? | 1-2 |
| Gender | 4 | Was the researcher male or female? | 8 |

| | | | |
|---|---|---|---|
| Experience and training | 5 | What experience or training did the researcher have? | Supplementary Table S1 |
| *Relationship with participants* | | | |
| Relationship established | 6 | Was a relationship established prior to study commencement? | Supplementary Information S2 |
| Participant knowledge of the interviewer | 7 | What did the participants know about the researcher? e.g. personal goals, reasons for doing the research | Supplementary Information S5 |
| Interviewer characteristics | 8 | What characteristics were reported about the inter viewer/facilitator? e.g. Bias, assumptions, reasons and interests in the research topic | 8 |
| **Domain 2: Study design** | | | |
| *Theoretical framework* | | | |
| Methodological orientation and Theory | 9 | What methodological orientation was stated to underpin the study? e.g. grounded theory, discourse analysis, ethnography, phenomenology, content analysis | 7 |
| *Participant selection* | | | |
| Sampling | 10 | How were participants selected? e.g. purposive, convenience, consecutive, snowball | 7 |

| | | | |
|---|---|---|---|
| Method of approach | 11 | How were participants approached? e.g. face-to-face, telephone, mail, email | 7 |
| Sample size | 12 | How many participants were in the study? | 11 |
| Non-participation | 13 | How many people refused to participate or dropped out? Reasons? | Supplementary Figure S3 |
| *Setting* | | | |
| Setting of data collection | 14 | Where was the data collected? e.g. home, clinic, workplace | 8 |
| Presence of non-participants | 15 | Was anyone else present besides the participants and researchers? | Supplementary Information S4 |
| Description of sample | 16 | What are the important characteristics of the sample? e.g. demographic data, date | 8;<br>11-12; |
| *Data collection* | | | |
| Interview guide | 17 | Were questions, prompts, guides provided by the authors? Was it pilot tested? | 7-8;<br>Supplementary Information S3 |
| Repeat interviews | 18 | Were repeat interviews carried out? If yes, how many? | Supplementary Information S2 |
| Audio/visual recording | 19 | Did the research use audio or visual recording to collect the data? | 8 |

| | | | |
|---|---|---|---|
| Field notes | 20 | Were field notes made during and/or after the interview or focus group? | 9 |
| Duration | 21 | What was the duration of the interviews or focus group? | 8-9 |
| Data saturation | 22 | Was data saturation discussed? | This study used iterative and responsive data collection, employing both focus groups and follow-on interviews. Additionally, this study was designed used co-design principles, and its results will inform future iterative rounds of data collection to refine understanding. As such, our sample size was determined based on data richness and adequacy to answer the research question and the ability to inform future co-design, rather than on concepts of data saturation.(1-3)<br><br>1. Braun V, Clarke V. To saturate or not to saturate? Questioning data saturation as a useful concept for thematic analysis and sample-size rationales. Qualitative Research in Sport, Exercise and Health. 2019;13:201 - 16.<br>2. Malterud K, Siersma VD, Guassora AD. Sample Size in Qualitative Interview Studies: Guided by Information Power. Qualitative Health Research. 2016;26(13):1753-60.<br>3. Saunders B, Sim J, Kingstone T, Baker S, Waterfield J, Bartlam B, et al. Saturation in qualitative research: exploring its conceptualization and operationalization. Quality & Quantity. 2018;52(4):1893-907. |

| | | | |
|---|---|---|---|
| Transcripts returned | 23 | Were transcripts returned to participants for comment and/or correction? | 9;<br>Supplementary Information S4 |
| **Domain 3: analysis and findings** | | | |
| *Data analysis* | | | |
| Number of data coders | 24 | How many data coders coded the data? | 9-10 |
| Description of the coding tree | 25 | Did authors provide a description of the coding tree? | Supplementary Table S2 |
| Derivation of themes | 26 | Were themes identified in advance or derived from the data? | 9-10 |
| Software | 27 | What software, if applicable, was used to manage the data? | 10 |
| Participant checking | 28 | Did participants provide feedback on the findings? | 9 |
| *Reporting* | | | |
| Quotations presented | 29 | Were participant quotations presented to illustrate the themes/findings? Was each quotation identified? e.g. participant number | 14-19;<br>Supplementary Table S8 |
| Data and findings consistent | 30 | Was there consistency between the data presented and the findings? | 14-19;<br>Supplementary Table S3, S7, S8 |
| Clarity of major themes | 31 | Were major themes clearly presented in the findings? | 14-19 |

| | | | |
|---|---|---|---|
| Clarity of minor themes | 32 | Is there a description of diverse cases or discussion of minor themes? | 14-19 |

Consolidated criteria for reporting qualitative research (COREQ): Tong A, Sainsbury P, Craig J. Consolidated criteria for reporting qualitative research (COREQ): a 32-item checklist for interviews and focus groups. Int J Qual Health Care. 2007;19(6):349-57.

### Supplementary Figure S3: Recruitment flowchart

*Supp. Figure S1: Study recruitment flowchart*

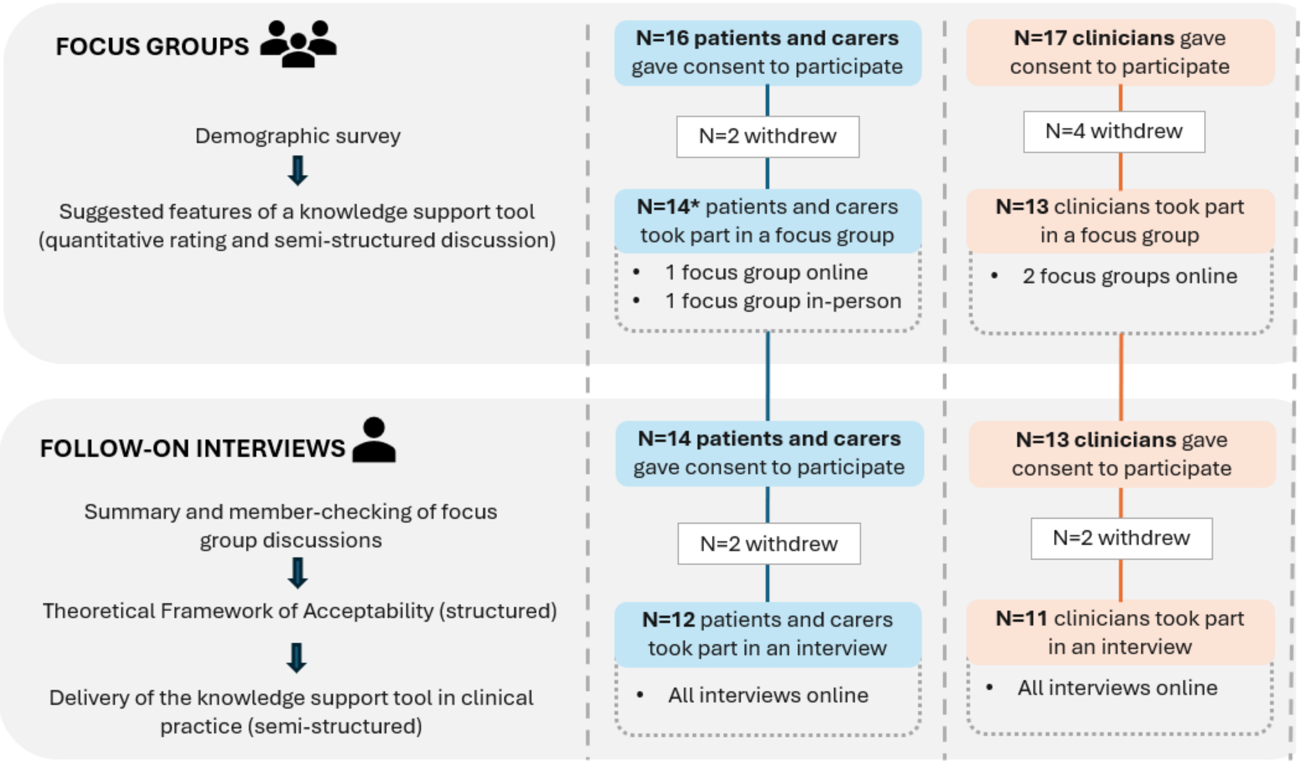


*One patient/carer participant attended both focus groups. The count reflects the total number of unique participants.

Clinician participants withdrew from focus groups due to no longer being able to attend a scheduled focus group date (N=4). Patient/carer participants withdrew from focus groups due to longer being able to attend a scheduled focus group date (N=2).

Clinician participants withdrew from interviews due to (1) lack of time to attend an interview (N=1), and (2) due to lack of response to interview invite (N=1). Patient/carer participants withdrew from interviews due to (1) lack of time to attend an interview (N=1), and (2) without reason given (N=1).

No repeat interviews were carried out.

### Supplementary Table S7: Utility ratings

*Table S7: Utility ratings for suggested content and functionality of a knowledge support tool: types and illustrative examples*

| Content/functionality type | Median (IQR) utility [N (%)] | | Example content/functionality | Median (IQR) utility [N (%)] | |
|---|---|---|---|---|---|
| | Patients/ carers* | General practice clinicians | | Patients/ carers* | General practice clinicians |
| Guidance about what to do during a GP consultation with someone who has self-harmed[2] | 10 (8-10) [N=16 (114.3%)] | - | Prompts to help assess risk of self-harm or suicide | 6 (4.5-8.5) [N=15 (107.1%)] | - |
| | | | Prompts to discuss individual care needs (including care plans) | 8 (6.5-9) [N=15 (107.1%)] | - |
| | | | Guidance about what treatment or assessments a patient should receive | 7 (5.5-9) [N=15 (107.1%)] | - |
| Information on best-practice guidelines (eg NICE) for self-harm | - | 7 (7-9) [N=13 (100%)] | Information from NICE guidelines about self-harm management/assessment and suicide prevention | - | 7 (5-8) [N=13 (100%)] |
| | | | Flagging when guideline recommendations have not been met (e.g. high-risk prescribing) | - | 8 (7-10) [N=13 (100%)] |

| | | | | | |
|---|---|---|---|---|---|
| | | | Information from studies/other published sources which may help guide self-harm management/assessment | - | 5 (3-6)<br><br>[N=13 (100%)] |
| Prompts to support communication and documentation of self-harm | - | 8 (6-9)<br><br>[N=13 (100%)] | Prompts to ensure self-harm is coded into the health record | - | 8 (5-9)<br><br>[N=13 (100%)] |
| | | | Prompts to assess presence of risk factors which may be associated with harmful outcomes (such as suicide) | - | 9 (8-10)<br><br>[N=13 (100%)] |
| | | | Prompts to discuss individual care needs (including care plans) | - | 8 (6-9)<br><br>[N=13 (100%)] |
| Information about a patient's medicines, care plans or clinical history | 6 (5-10)<br><br>[N=15 (107.1%)] | 9 (5-10)<br><br>[N=13 (100%)] | A summary of medication prescribing history | 8 (7.5-10)<br><br>[N=15 (107.1%)] | 10 (8.5-10)<br><br>[N=12 (92.3%)] |
| | | | A summary of clinical events history (e.g. diagnoses)† | 5.5 (4.2-9.8)<br><br>[N=15 (107.1%)] | 10 (7-10)<br><br>[N=12 (92.3%)] |
| | | | Services that have been previously or currently accessed (e.g. hospital visits and referrals) | 8<br>(6-9.5) | 10<br>(8-10) |

| | | | | | |
|---|---|---|---|---|---|
| | | | | [N=15 (107.1%)] | [N=12 (92.3%)] |
| Information on support that may be available outside of GP appointments | 10 (8.8-10) [N=16 (114.3%)] | 10 (9-10) [N=13 (100%)] | NHS referral pathways | 8 (6-10) [N=15 (107.1%)] | 9 (8-10) [N=11 (84.6%)] |
| | | | Community or third-sector support (regional or national) | 10 (8.5-10) [N=15 (107.1%)] | 9 (8.5-10) [N=11 (84.6%)] |
| | | | Helplines | 6 (5-8.5) [N=15 (107.1%)] | 9 (9-10) [N=11 (84.6%)] |
| The ability to provide better monitoring of patients outside of GP appointments | 5.5 (4-6.8) [N=15 (107.1%)] | 5 (5-6) [N=13 (100%)] | Surveys or apps that can be used to log mood, thoughts related to self-harm and suicidal ideation | 3 (1.2-4.5) [N=15 (107.1%)] | 5 (2.5-5) [N=11 (84.6%)] |
| Information leaflets for patients/carers to refer to after leaving the GP practice | 7.5 (5.2-8.8) [N=15 (107.1%)] | 9 (9-10) [N=13 (100%)] | Information leaflets that explain what self-harm is or give advice on how to manage self-harm | 6.5 (3.5-8.8) [N=15 (107.1%)] | 9 (7.8-10) [N=12 (92.3%)] |
| | | | Information leaflets that explain what medicines a patient is taking, or care they are receiving | 8 (6.5-8.5) [N=15 (107.1%)] | 9 (8.5-10) [N=12 (92.3%)] |

| | | | | | |
|---|---|---|---|---|---|
| | | | Information leaflets that give details for available support outside of GP appointments | 8 (7.5-10)<br><br>[N=15 (107.1%)] | 10 (9-10)<br><br>[N=12 (92.3%)] |
| | | | Information leaflets in accessible/easy-read formats and different languages | 10 (9-10)<br><br>[N=15 (107.1%)] | 10 (10-10)<br><br>[N=12 (92.3%)] |

IQR=interquartile range, N=number of participants, NICE=National Institute for Health and Care Excellence, GP=general practice.

Median values, their IQRs and percentages were reported to one decimal point.

Utility was ranked on a scale of 1-10, with 1=completely not useful, 5=neutral, and 10=very useful.

'Information from guidelines' and 'Prompts to support communication and documentation' were presented only to clinicians, as these are both relate to healthcare professional facing content (e.g. it was not expected that patients or carers would use or be familiar with clinical guidelines). Patients/carers were instead presented with the broader option, 'Guidance for clinicians on what to do during a consultation', which was intended to broadly capture both best-practice guidance and support for communication. As such, direct comparisons between patients/carers and clinicians were not possible for these items.

*Some patient/carer participants submitted more than one response (one participant attended both focus groups). All responses were provided anonymously and therefore multiple responses could not be identified.

[†]Patients/carers were instead presented with 'summary of conditions a patient has been diagnosed with'; this narrower scope was intended to reduce conceptual breadth and increase clarity.

### Supplementary Table S8: Further illustrative quotes

*Table S8: Illustrative quotes for the qualitative themes*

| |
|---|
| Theme 1: Content to facilitate person-centred care |
| Subtheme 1: Building informational and management continuity |
| Quote 1, C_FG13, focus group[1]: *“The number of patients that say they've tried this, they've tried that, they've had side effects to such a body, and you have to trawl through the records to find out what’s gone on in the past, especially if they go from one doctor to another. So, having a summary of when they were prescribed as a snapshot would be really helpful… I suppose how long is a piece of string. But going back for the last two years even, just to know if they've kept coming and changing it”*<br><br>Quote 2, C_I5, interview: *“I do wonder if it would struggle with really complex presentations… a lot of drug use, vulnerabilities, homelessness, no digital access… I think it might struggle to assimilate all the information for those.”*<br><br>Quote 3, PC_FG14, focus group: *“You might be receiving treatment from an out of locality area and that is your main source of treatment but if you haven’t seen your GP for the last ten months, they’re not going to have any information. That summary is going to be empty… there needs to be a way that all that information can be incorporated because otherwise, what is the summary going to have because some people don’t go and visit their GP, it’s really hard to get a GP appointment but they will access other services in the community.”*<br><br>Quote 4, PC_FG1, focus group[1]: *“Say you’ve gone three times and one of those times [a support option] has already been suggested to you, could it come up as part of a tool like ‘it’s been suggested previously’ so you’re not going over old ground… It could potentially be linked into the whole tracking of progress thing as well.”*<br><br>Quote 5, PC_FG2, focus group: *“A lot of emphasis does get put on the helplines. The helplines are good but it really depends on who you get on the other end. I’ve had some really good experiences, but then I’ve had some really awful experiences with helplines. I know a lot of time when I get given, have you got the helpline numbers, I sigh and go yes. It always seems to be the first thing that’s asked.”* |
| Subtheme 2: Responsive to patients’ choice and control |

| |
|---|
| Quote 6, PC_FG3, focus group: *“There are certain things that I don’t want everybody knowing. I’ve got a misdiagnosis on my medical record, and I know that as soon as somebody sees that they’re going to say ‘oh no, not again’, and it’s just rubbish”*<br><br>Quote 7, PC_FG4, focus group[1]: *“I have a physical disability, but quite often if they see mental health they’ll associate the two. And if it’s a mental health appointment often it gets blamed on the physical health, which obviously does play an element, but more the other way round that if it was physical health and they saw mental health there’s quite assumption that you’re not actually physically disabled, you’re mentally ill. That link is so common that it makes me nervous that if I went in for something specific and they had it all up on a screen that would be the first impression”*<br><br>Quote 8, PC_FG14, focus group[1]: *“Sometimes your physical health will take absolutely precedence over your mental health, even though the two are completely inter-related”*<br><br>Quote 9, PC_FG6, focus group: *“I’m disabled with chronic illness, and I’d be really cautious about who that information was being shared with and for what purpose. If I have a good stretch with my illnesses is that going to end up in the hands of the DWP who are going to say, you’re not really that poorly, are you? Do you know what I mean?”*<br><br>Quote 10, C_FG4: *“We can't have hundreds of potential communications from patients between consultations. That said, I do think that it would be really useful an hour before a prebooked appointment to have this information [about patients’ mood, feelings and thoughts] … in the same way as we do home blood pressure monitoring where they measure blood pressure over a period and then we get that summary. Absolutely, that could be a really useful thing for patients to do.”* |
| Subtheme 3: Supporting holistic care: opportunities and tensions |
| Quote 11, PC_I3, interview[1]: *“The tool could prompt the GPs to ask certain questions. ‘Would you like to change something about what you do?’, ‘how do you feel about your self-harm? Is it something that you’ve accepted, something you like, something you don’t like?’, ‘do you feel has got worse and this is why you're here today?’ Especially if somebody’s been self-harming for a long time, why did they decide to go to the GP?”*<br><br>Quote 12, PC_FG4, focus group: *“Sometimes I wish there was more guidance. Because I’ve looked at my notes on the NHS app before and it’s made a comment and been like, ‘looks put together’, or something like that. And it’s like, okay, but if that’s contributing to your risk assessment of me that’s such a superficial way of assessing my risk.”*<br><br>Quote 13, C_I2, interview: *“[Questions used to assess risk] varies depending on who they [the patient] are and how it’s going, sometimes they will volunteer quite a lot of that information anyway …Have they had psychosis before? Then they are more likely to be risky, very isolated. Are they quite impulsive, do they gamble, do drugs, do lots of alcohol? Who have they got around them, any mates, family?* |

| |
|---|
| *Geographically and socially as well. Then what they tend to do when they are in a moment of crisis. Do they isolate themselves, do they head home? I won't necessarily ask all of those questions in one but just based on the person in front of me, I'll tailor some of those"* |
| Quote 14, C_I9, interview[1]: *"It will be really useful in my role for risk stratification, especially for allocation of appointments… We have a mental health care coordinator and ultimately we'd love her to be able to decide the appointments without us… We've looked to develop a scoring sort of thing that a care coordinator could - because she's not medical, that's where it keeps falling down because somebody always inevitably says 'well, the risk's too great'. You're asking someone who's not clinical to make the decision. But if a tool could make the decision that might help"* |
| Quote 15, C_I2, interview: *"If this tool would have a different grade of the current level of self-harmingness, like the PHQ score, so we can look at the trend of it."* |
| Quote 16, C_FG9, focus group[1]: *"[Risk scores] lose the human humanistic side of what we're trying to achieve… We've got patients who are severely mentally ill, but they don't fulfil the criteria to require admission. But it affects their daily life in such a tremendously significant way, and yet the tool, the book says, no, you don't qualify. But as a GP, which is the nice thing about being a GP, you try to be holistic, you try to take the whole person. And [a score] could potentially just strip somebody of that personification. It can just give them a number, score this from one to ten."* |
| Quote 17, C_I2, interview: *"My surgery deals with a lot of patients from the BAME cohort where self-harm is considered taboo… In some cases, maybe adding a bit of culture would be more beneficial for them, religion would be more beneficial for them because religion is one aspect which is a deterrent for self-harm"* |
| Quote 18, PC_FG8, focus group: *"The most recent issues I had were hormonal. So, it wouldn't have come under mental health, so it would have got left out with that filter. So, filtering sometimes is useful but sometimes it could leave a lot of stuff that they actually need out."* |
| Quote 19, C_FG13, focus group: *"It would be really useful for, one, being patient specific, so if it has knowledge of the patient, so not just about the self-harm but what's led that person to self-harm. So, you know, if it's an asylum seeker, if it's a young person, domestic abuse, gambling …so it's not just necessarily available support for the self-harm, but the other aspects of the consultation."* |
| Theme 2: Accessibility and usability |
| Subtheme 1: Practical and actionable |
| Quote 20, C_I1, interview[1]: *"Having something that's pre-made on Accurx that you can copy and paste or have an automated template that you can click and send to patients I think's really powerful… I think sometimes it can be helpful to give a follow-up. I think it's more* |

| |
|---|
| *about if you developed these thoughts, these are your options in terms of support… Practical information which you can share with patients so that they've got safety netting… Having a documented plan for if things were to deteriorate is quite helpful from a medicolegal point of view as much as anything else."* <br> Quote 21, PC_FG8, focus group: *"The NICE guidelines are quite general and common sense and more about just reminding people that they're supposed to be compassionate, that they're supposed to treat the person as an individual with respect. But apart from that, it doesn't really give them the practical stuff that they're looking for. So, it wouldn't really tell what you should really say, at this moment, what can you suggest to this person."* <br> Quote 22, PC_FG1, focus group: *"I think it would be really useful basic first aid information for me. Some people literally have no idea, 'I don't know when I need to go and get stitches', or 'I don't know if this is a sign of infection or if I'm just overreacting'. So, I think basic information around signs of infection, when you need to get a cut looked at, just stuff around burns maybe, when a burn needs to be checked out, I think that is really good knowledge for anybody who self-harms or supports anyone who self-harms to have."* <br> Quote 23, PC_FG4, focus group: *"I think sometimes there needs to be slightly clearer or more direct questions. Because sometimes I'm not just going to come out and say it because that's intense, but if someone asks you, you might be more likely to say it and try and get help."* <br> Quote 24, PC_FG1, focus group: *"I think these resources definitely need to be user friendly and coproduced to make them useful at all. Intimidating language only makes things more inaccessible"* <br> Quote 25, PC_FG8, focus group[1]: *"The prompts are supposed to be to engage risk but also to start building a relationship and building trust. It's not about ticking boxes, are we trying to cover our backs or are they actually really trying to get through to this person?"* <br> Quote 26, C_FG4, focus group: *"It's important to document risk, but actually it can be very, very powerful to document verbatim what patients say. And sometimes we can lose that on some of these tick boxing"* |
| Subtheme 2: Streamlining workflows |

| |
|---|
| Quote 27, C_FG1, focus group[1]: *"If it's screeds of text, that's useless in the consultation. I'm sure you've come across Ardens. Often their tools are dynamic, so if you tick boxes, it then tells you what to do. That kind of thing is really helpful. Whereas if it's just a link to the NICE guidelines, I'd never click on it."*<br>Quote 28, PC_I2, interview: *"Maybe just a reminder, even if it was five bullet points or an acronym. Something that people could just be like, 'oh yeah, this is what I need to remember'. Then obviously you'll need more in-depth stuff and it wouldn't be case-specific. I just feel like that could be helpful if you're short on time."*<br>Quote 29, C_FG5, focus group: *"If there's anything that aids quicker documentation and coding, yes, I'm all for that."*<br>Quote 30, C_FG4, focus group: *"Recreating something is not necessarily necessary 'cause there are fantastic resources already available. But, yeah, accessing them or finding them in the context of a consultation so you'd have them handy is actually the more useful thing."*<br>Quote 31, C_FG12, focus group: *"A referral might be helpful, and now I'm going to have to go away and do the letter or whatever's required. But if the decision support tool has already identified that itself, and says, do you agree with me, I've already prepared the referral for you, would you like me to process it, then that'd be great. 'Cause you can do that within the appointment. You can just say, yes, and move on."*<br>Quote 32, C_FG6, focus group: *"We've set this up as a preset Accurx on our system, so we have that information to provide. It's just one click and it's gone, so it's just about making this as slick and integrated to what we do as possible"*<br>Quote 33, C_FG12, focus group: *"If there's an opportunity to skew the tool in that direction [of summarising the electronic health record], I'd find that more useful than the management suggestion side of it. Perhaps other clinicians or allied health professionals/nurses might find it useful to have more support in terms of the clinical decision making but if I had all the information there to me, available in a click, I think I'd feel very happy just to go ahead and make my own decision"* |
| Theme 3: Balancing competing priorities of general practice |
| Quote 34, C_FG9, focus group: *"I don't like to be the centre of attention in that I'll have some responsibility but I think with the cohort of patients we're talking about, there are other professionals and other services that are involved. I wouldn't want the treatment, the management, to be centred and focused on GP."* |

Quote 35, C_FG13, focus group: *“If a patient’s self-reporting outside of the hours of the GP opening and if they're putting in things like, I'm terrible today, I'm going to self-harm, it’s how is that notified. And then once you've seen it, how do you respond and your clinical responsibility, because again, I'm slightly cynical but sometimes our patients try to access because they obviously want that attention”*

Quote 36, C_FG1, focus group: *“[Summarising information about a] care plan scares me because care plans are really, really long sometimes, so it’s how that’s presented. You know, fine, press a button to view the care plans if you want, but it’s just getting the key information in a quick way... what might be more useful would be to give patients tools to deliver their own care plan”*

Quote 37, C_FG9, focus group[1]: *"I think any application that...not passes the buck, but essentially encourages self-care. More and more we try to signpost patients/customers to the right place. And [providing information about support services] is quite a useful one, ‘cause once it hits my desk there’s accountability and responsibility and all that stuff that you find it...there’s a bias there and you have to dispose – using that word very loosely – of that patient. You have to. Whilst if you give them the option to look outside this GP service, first of all, so we do know what’s out there but it’s no use to us unless it can be directed to the user."*

Quote 38, PC_I3, interview: *“At the end of the day you want the patient to feel they got listened to and if they managed to walk away with something different that would be a fantastic result. But being listened to is going to be the key.”*

Quote 39, PC_I3, interview: “*At the end of the day it will be two people, so there’s two attitudes that’s going to be involved in this, and the GP might be willing and the person goes in... all guns blazing, wanting you to fix them on the spot, which you can't do. So I guess, again the two people play quite a big part in this. Just trying to think, my interactions with GPs, and especially when I first reached out, I think that GP could have really used this. It was quite obvious that he didn’t have a clue what to say and what to do. So he chose to say nothing and do nothing. I wasn’t expecting to be fixed, I was just looking for some guidance and he didn’t have a clue.”*

Quote 40, PC_I3, interview: *“It could be very different when somebody goes after the first instance of self-harm or someone who goes after 20 years of self-harming. What their expectations is going to be and what the help is going to be is going to look very different”*

Quote 41, PC_FG1, focus group: *“We all know how limited GP’s appointments are and limited the amount of support you can actually get. So, if I went to the GP I wouldn’t be expecting having ongoing support from that person, so to be directed to some other service would feel useful for me”*

Quote 42, PC_FG8, focus group[1]*: “Maybe some [guidance around expectations] would be useful in the fact that this isn’t about waving a magic wand and self-harm will disappear because that just won’t happen. So, how can they also help the person who has limited expectations of themselves and of the services. From what I’ve seen, it’s more like, ‘oh my god, oh my god, I better not panic in front of this person... let’s just quickly find something to maybe suggest, to just get them out the door and hopefully next time they will come to*

| |
|---|
| *see a different one and not me'. In some cases what might be more useful is what they're asking the person, how they are engaging in that conversation with a patient, not necessarily coming up with solutions because that could make a massive difference in somebody's life instead of, oh they gave me the leaflet or they've sent me back, they've asked me about the thing that I already know."* |

Anonymous identifier key: PC=patient/carer, C=clinician, FG=focus group, I=interview.

Focus group data were anonymised prior to interviews, and therefore data from the same participant cannot be linked across focus groups and interviews.

[1]Quote is included in-part in the main text

### Supplementary Information S5: Ethics

The study was conducted in conformance with the Declaration of Helsinki, Good Clinical Practice and the UK Policy Framework for Health and Social Care Research. Prior to both the focus group and interview, participants were given a detailed study information sheet and consent form and provided either written or verbal consent. The study information sheet which outlined the rationale for this study, who will conduct the research, when it will take place, how the data will be used, how the data will be stored, details about anonymity and confidentiality, and how participants can choose to withdraw. Consent was considered an ongoing process, with withdrawal allowed at any time prior to anonymisation. Data were anonymised as soon as possible following transcription, which included the removal of any potentially identifiable information and deletion of recordings. As such, focus group data were anonymised prior to interviews, and cross-data linkage was not possible. Data were stored on a secure server in-line with data protection standards.
